# Surgery-related obstetric fistulas across health facilities in sub-Saharan African countries: systematic review and meta-analysis

**DOI:** 10.1101/2025.09.25.25336674

**Authors:** Mercy M. Imakando, Ernest Maya, David Owiredu, Mercy W. Monde, Choolwe Jacobs, Kwadwo O. Akuffo, Isaac Fwemba, Anthony Danso-Appiah

## Abstract

**Background:** Surgery-related (iatrogenic) obstetric fistulas are a major public health threat in sub-Saharan Africa (SSA), but the actual burden is largely unknown due to a paucity of data. This review aimed to collate empirical evidence on the magnitude and determinants of iatrogenic obstetric fistulas in SSA.

**Methods:** PubMed, LILACS, CINAHL, SCOPUS and Google Scholar were searched from 1^st^ January 2000 to 31^st^ March 2025, without language restrictions, via comprehensive search terms. The Cochrane Library, African Journals Online, Data Base of African Thesis and Dissertations, including Research (DATAD-R D Space), Preprint Repositories and reference lists of relevant studies, were also searched. The retrieved studies were deduplicated in Endnote, screened and selected in Rayyan. Two authors independently selected studies, extracted data and assessed the quality of the included studies via pretested tools. Disagreements between reviewers were resolved through discussion. Weighted proportions were estimated via STATA SE 18. Heterogeneity between studies was assessed graphically and statistically, and where significant, random effects model meta-analysis was performed. Estimates are reported with 95% confidence intervals (CIs). Subgroup analysis was conducted to address heterogeneity, and sensitivity analyses were performed to test the robustness of the pooled estimates.

**Results:** The proportion of surgery-related obstetric fistulas among urogenital/rectovaginal fistulas in SSA is 40% (95% CI 34–47, 60 studies, n=25825), whereas among obstetric fistulas, it is 44% (95% CI 37–51, 55 studies, n=17504). The procedures that frequently preceded obstetric fistulas were as follows: caesarean section (34%, 95% CI 28–40; 54 studies, n=17491) and instrumental vaginal deliveries (11%, 95% CI 9–13; 34 studies, n=12727). Most procedures that resulted in obstetric fistulas were performed by nonspecialists (92%, 3 studies, n=749).

**Conclusions:** The proportions of iatrogenic obstetric fistulas among urogenital/rectovaginal fistulas (40%) and obstetric fistulas (44%) are high and raise serious concerns about the quality of obstetric surgical care delivery in SSA. The fact that more than 92% of obstetric surgeries are performed by nonspecialists and that the incidence of iatrogenic fistulas is high calls for increased efforts towards capacity building and quality improvement across countries in SSA.

**Registration:** The protocol for this systematic review was registered in the International Prospective Register for Systematic Reviews (PROSPERO) with registration ID: CRD42021277993.

## Background

Obstetric fistulas are serious injuries sustained by mothers during childbirth, that lead to abnormal connection(s) between the vagina and the urinary tract (urethra, bladder or rarely ureters) or the rectum, that can result in urine and/or faecal incontinence [1, 2]. Globally, over 2 million women of reproductive age suffer from obstetric fistula, with thousands of new cases every year [3]. Most of these cases occur in sub-Saharan Africa (SSA) and Asia [4] due to weak health systems and failure to provide universally accessible, timely and quality obstetric care [5]. Most high income countries in Europe and the United States of America (USA) have been able to address fistula development as way back as the 1950s as a result of availability and access to quality healthcare delivery and emergency obstetric care (EmOC) services [6]. In contrast, SSA still struggles with a shortage of trained staff, a lack of or limited medical supplies, poor quality of care, long waiting times, poor referral systems and poor coordination of tasks among staff [7]. The long distance pregnant women have to travel to access the nearest health facility, poor road networks and high transport costs further limit pregnant women’s access to quality care during childbirth across countries in SSA [8, 9].

The main cause of obstetric fistulas is obstructed labour [10–15] and complications resulting from surgery (i.e. iatrogenic fistulas) [16]. Other determinants of obstetric fistulas are small maternal pelvis, foetal mal-presentations or abnormalities, and therapeutic misadventure [17, 18]. Early child bearing [9, 19], low educational levels [20–22] and poverty [23], as well as cultural factors such as female genital mutilation [24] and the patriarchal systems in SSA, also contribute to the high occurrence of obstetric fistula [25].

Attempts to increase access to EmOC and reduce maternal morbidity and mortality has resulted in an increase in caesarean section (CS) rates [26], which in turn has led to more iatrogenic obstetric fistulas [27]. Iatrogenic fistulas have also been increasing in cases of elective CS [28]. This raises serious concerns about the quality of obstetric care in SSA [29] and the training needs of healthcare professionals across SSA [30]. A multi-country study involving SSA countries reported that over 90% of obstetric surgeries resulting in fistulas were performed by nonspecialists [28]. This has been attributed to the widespread surgical task shifting commonly employed in low- and middle-income countries (LMICs) in attempts to mitigate the critical shortage of the specialist surgical workforce [31–33]. Currently, there are fewer than three specialists per 100,000 population in most SSA countries [34], of whom only a fraction are qualified obstetricians.

Addressing iatrogenic obstetric fistula is an integral component of Sustainable Development Goals (SDGs) 3 and 5.6—ensuring healthy lives for all and promoting universal access to sexual and reproductive health by 2030 [5]. Therefore, it is imperative that urgent effort be directed towards reducing and/or preventing this tragic complication of childbirth [35]. This systematic review is not duplicating existing reviews (see statements about this in the published protocol) [36] and sought to determine the burden of surgery-related obstetric fistulas occurring in countries across SSA and the type and experience of personnel (specialists or nonspecialists) performing surgical operations. The findings and conclusions will help inform sound policies, programs and human resource planning aimed at addressing the increasing levels of iatrogenic fistulas in SSA. Further details of the introduction can be found in the published protocol [36] https://pmc.ncbi.nlm.nih.gov/articles/PMC11346637/pdf/pone.0302529.pdf.

## Methods

The methods of this systematic review has been reproduced from the published protocol [36], which followed guidelines specified in the Cochrane Handbook [37] and formats of previously published reviews [38–43]. It been reported in line with the Preferred Reporting Items for Systematic Review and Meta-Analysis (PRISMA) guidelines [44]. The study retrieval and flow through the selection process are summarized in the PRISMA flow diagram [45]. The protocol for this systematic review and meta-analysis was registered in the International Prospective Register for Systematic Reviews (PROSPERO), with registration ID CRD42021277993.

### Criteria for considering studies for this systematic review

#### Types of studies

Informed by the published protocol [36] and earlier published works [38–41], it included randomized controlled trials (RCTs), quasi-RCTs, cohort studies, case‒control studies and cross-sectional studies were eligible for inclusion in this review. Case studies and case series were not eligible for inclusion because they are atypical and not representative of the source population. Studies using secondary data, commentaries, editorials, opinions and country-level statistical reports were not eligible for inclusion. This study did not incorporate reviews, as the unit of analysis was restricted to primary studies. Nevertheless, the reviews were carefully examined to identify any potentially eligible studies not captured in our searches. Global reviews that included SSA as a subset or subregional focus as a whole were not eligible; we included only the studies conducted in SSA. Studies that reported a country or regional estimate without a well-defined representative sample or subsample within the source population were not eligible for inclusion. For multi-country studies that included studies from SSA and reported data separately for each country, data from SSA countries were included. In cases where the results were combined with no way of disaggregating the data, the studies were excluded. Commentaries and opinions were not eligible for inclusion.

#### Types of participants

The review included women living in sub-Saharan Africa (SSA) with urogenital or rectovaginal fistulas resulting from complications of childbirth associated with surgical procedures, symphysiotomies, episiotomies, operative vaginal deliveries (forceps or vacuum delivery), caesarean delivery, caesarean hysterectomy or laparotomy due to a ruptured uterus. Women with urogenital or rectovaginal fistulae resulting from normal vaginal delivery, assault, gynaecological surgeries, malignancies or radiation were not considered, except when reported concurrently with obstetric fistulas.

#### Intervention/exposure

The exposure of interest was obstetric surgery, including CS, laparotomy for repair of the ruptured uterus, subtotal hysterectomy or hysterectomy for obstetric reasons such as a ruptured uterus and intractable postpartum haemorrhage.

#### Controls

The controls consisted of women with urogenital or rectovaginal fistulas resulting from vaginal deliveries without any obstetric surgery.

#### Outcomes

##### Primary outcome

- The proportion of iatrogenic obstetric fistulas among obstetric fistula patients was calculated as follows:

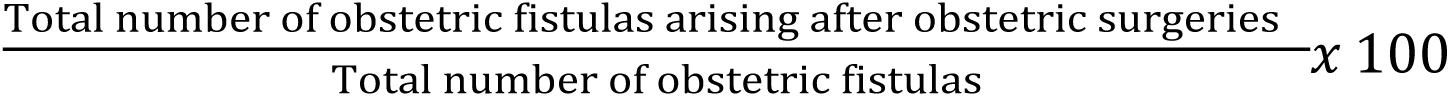
- The proportion of iatrogenic obstetric fistulas among genitourinary/rectovaginal fistula surgeries was calculated as follows:

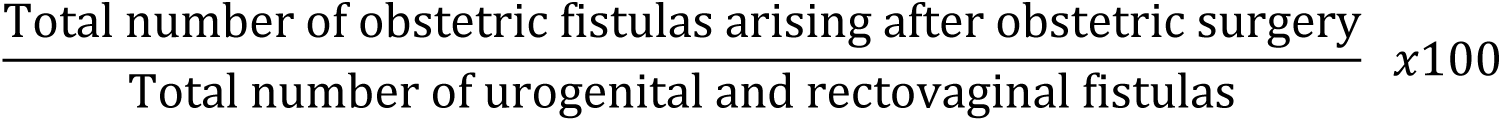

##### Secondary outcomes

- Personnel performing obstetric surgeries, i.e., Clinical Officers/Medical Licentiates, General Medical Doctors/Practitioners, Registrars and Specialists Obstetrician-Gynaecologists
- Correlates of surgery-related obstetric fistulas among women in SSA

#### Search methods for the identification of studies

The search strategy and methods were reproduced from the published protocol [36]. Studies (published and unpublished) from a wide range of sources (database and non-database sources) were retrieved and assessed for eligibility against pre-specified eligibility criteria. The following databases: PubMed, CINAHL, Google Scholar, LILACS, and SCOPUS were searched from January 1, 2000 to March 31, 2025, without language restrictions, using search terms developed from the inclusion/exclusion criteria. We also ran searches on African Journals Online (AJOL), the Cochrane Library, and the Database of African Thesis and Dissertations, including Research (DATAD-R D Space) and preprint repositories for additional studies. The reference lists of all potentially relevant studies were checked to retrieve studies missed by our searches. Some experts in the field across Africa and those affiliated with the International Society of Obstetric Fistula Surgeons (ISOF) and the International Federation of Gastroenterologists and Obstetricians (FIGO) were contacted via email for any knowledge about unpublished studies that could be included in the systematic review.

#### Managing the search output and selecting studies

The results from the databases were exported to Endnote, and duplicate records were removed. The deduplicated studies were exported to Rayyan software [46] for screening and selection via a pretested study selection flowchart adapted from earlier reviews [43, 47]. Three authors independently screened the titles and abstracts of the retrieved articles and selected studies. The full texts of all potentially eligible studies were retrieved and assessed for inclusion. The flow of studies throughout the selection process is presented in the PRISMA flow diagram [36]. Disagreements were resolved through discussion between the review authors.

#### Assessment of the quality of the included studies

Assessment of the risk of bias in the included studies followed the published protocol [36]. All the included studies used observational design and were assessed for risk of bias using the quality assessment tool developed by Hoy et al. [48]. The tool assesses 10 domains, namely, representation, sampling, random selection, nonresponse bias, data collection, case definition, reliability tool, prevalence period, numerators and denominators. The first four domains assess the external validity in the included studies, whereas the remaining domains (5 – 10) assess internal validity. Responses to each of the 10 criteria on the tool were judged as ‘low’, ‘high’ or ‘unclear’ risk of bias. A positive response to the risk of bias item was graded as high risk, whereas a negative response was graded as low risk. If the response to the risk of bias item was unclear or not indicated, it was deemed an unclear risk of bias. Two reviewers independently assessed the quality of the included studies, and any differences in responses were resolved through discussion and, where necessary, with the help of a third reviewer. The overall rating of risk of bias was mainly based on the internal validity domains and was rated as ‘low’ or ‘high’ risk of bias.

#### Data extraction and management

The following data were extracted independently by two reviewers using pretested data extraction sheet developed in Excel: study ID, country of origin, year of study, study design, method of data collection, eligibility criteria and sample size. We also extracted data on criteria for diagnosis, types of fistulas, cause of fistulas, mode of delivery and foetal outcomes in antecedent pregnancy, number of iatrogenic obstetric fistulas, total number of obstetric fistulas and genitourinary/rectovaginal fistulas. Health system variables include the level of expertise of the operating surgeon (nonspecialists: Clinical Officer, Medical Officer, General Practitioner/General Medical Officers; and specialists: Registrar(s) or Senior Registrar/Consultant Obstetrician Gynaecologists). Any discrepancies were resolved through discussion. A more senior reviewer was engaged in resolving complex issues.

#### Data synthesis and assessment of heterogeneity

Following what has been reported in the protocol [36], Stata SE18 was used to run the statistical analyses, reporting pooled proportions and their 95% CIs. Heterogeneity was assessed at three levels: clinical, methodological, and statistical heterogeneity. For the assessment of clinical heterogeneity, we explored differences in study characteristics, such as the study populations, interventions, and outcomes. Methodological heterogeneity was used to explore differences between studies in terms of their design and quality dimensions, whereas statistical heterogeneity was used to assess the variation in effects between studies by inspecting the forest plots for overlapping CIs and the I^2^-squared statistic. Sources explored for the presence of heterogeneity in this systematic review included study designs and the country where the study was conducted, as it was anticipated that there would be differences in systems and resources (human and material) in different countries and settings. The countries were further grouped according to the United Nations geographic regions, namely, Eastern, Western, Middle and Southern Africa [49]. Additionally, differences in fistula type, type of personnel conducting the surgery and type of surgery prior to obstetric fistula development were explored. The identified sources of heterogeneity formed the basis for subgroup analyses. The I² statistic [50] was used to measure the extent of heterogeneity across the studies in the meta-analysis. Studies were considered to have a low level of heterogeneity if I^2^ was ≤ 25%, moderate heterogeneity when I² was 26–50%, and a high level of heterogeneity if I² was > 50% [51, 52]. We employed a weighted random-effects model for the analysis [53].

#### Dealing with missing data

We did not impute when addressing missing data but instead contacted primary study authors and asked for the raw data to enable us to extract the missing information. When it was not possible to obtain missing data, only records with complete data on the outcome were included, i.e., complete case analysis.

## Results

In total, 1758 articles were retrieved from various databases and non-database sources, 157 duplicates were removed, 1510 were not eligible, 91 full-text articles were assessed for eligibility, 63 met our inclusion criteria, and 60 studies were included in the analysis (Fig 1). Two articles [54, 55] were published from one study [54], and <u>the</u> data from three studies were subsets of other studies: Browning [56] was a subset of data from Browning and Whiteside [57] Ezegwui and Nwogu-ijoko [58] was a subset of Obi et al. [59], while most data from Ijaiya & Aboyeji [60] had been reported in an earlier study by the same authors [<u>61</u>. Twenty-five potentially relevant studies were excluded for the following reasons: wrong focus (20 studies) [14, 62–80], lack of a well-defined denominator (two studies) [81, 82], not a primary study (one study) <u>83</u>], wrong setting (one study) [84] and insufficient information (one study) [85].

**Fig 1.**
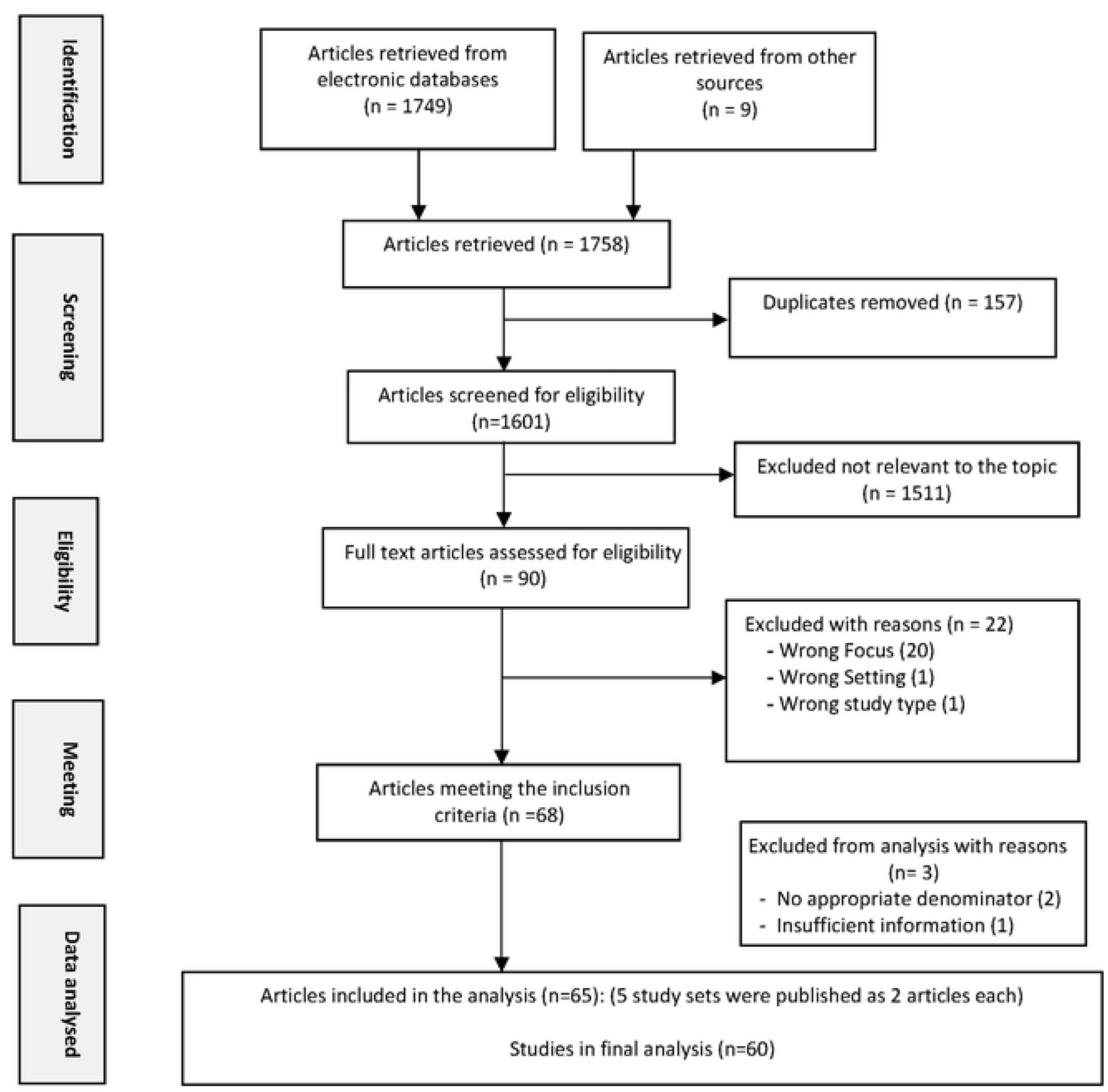
PRISMA flow diagram showing the study retrieval and selection process.

### Description of studies

The characteristics of the included studies are reported in Table 1. The studies included in this systematic review were conducted between 1988 and 2022 and were published between 2002 and 2024. Fifty-eight of the 60 included studies (96.7%) were published after 2003, the year the global campaign to End Fistulas was launched by the United Nations Fund for Population Activities (UNFPA) and its partners. In total, 25825 women with genito-urinary/rectovaginal fistulas, of whom 18707 had obstetric fistulas, were included in this systematic review.

**Table 1.**
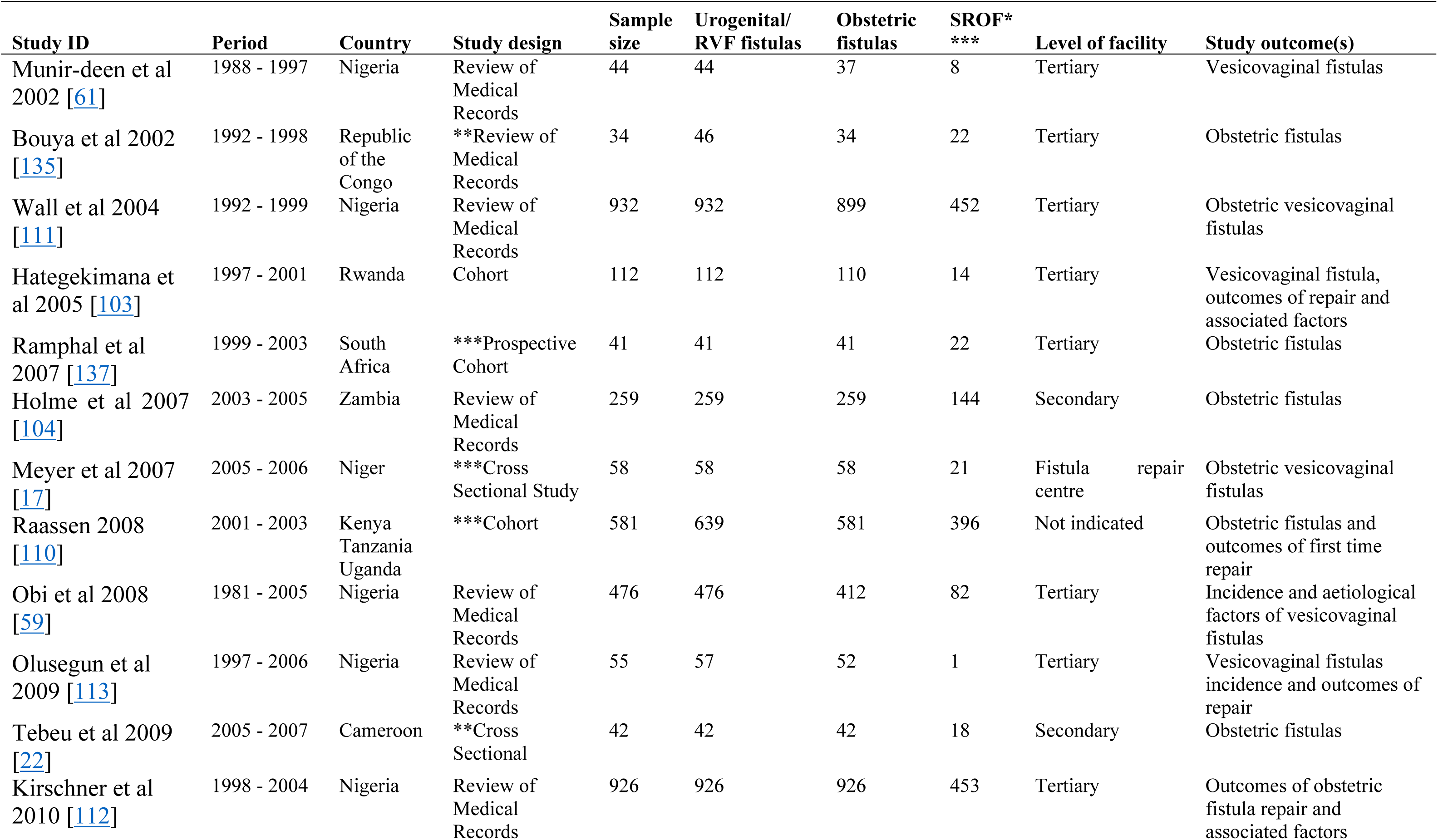
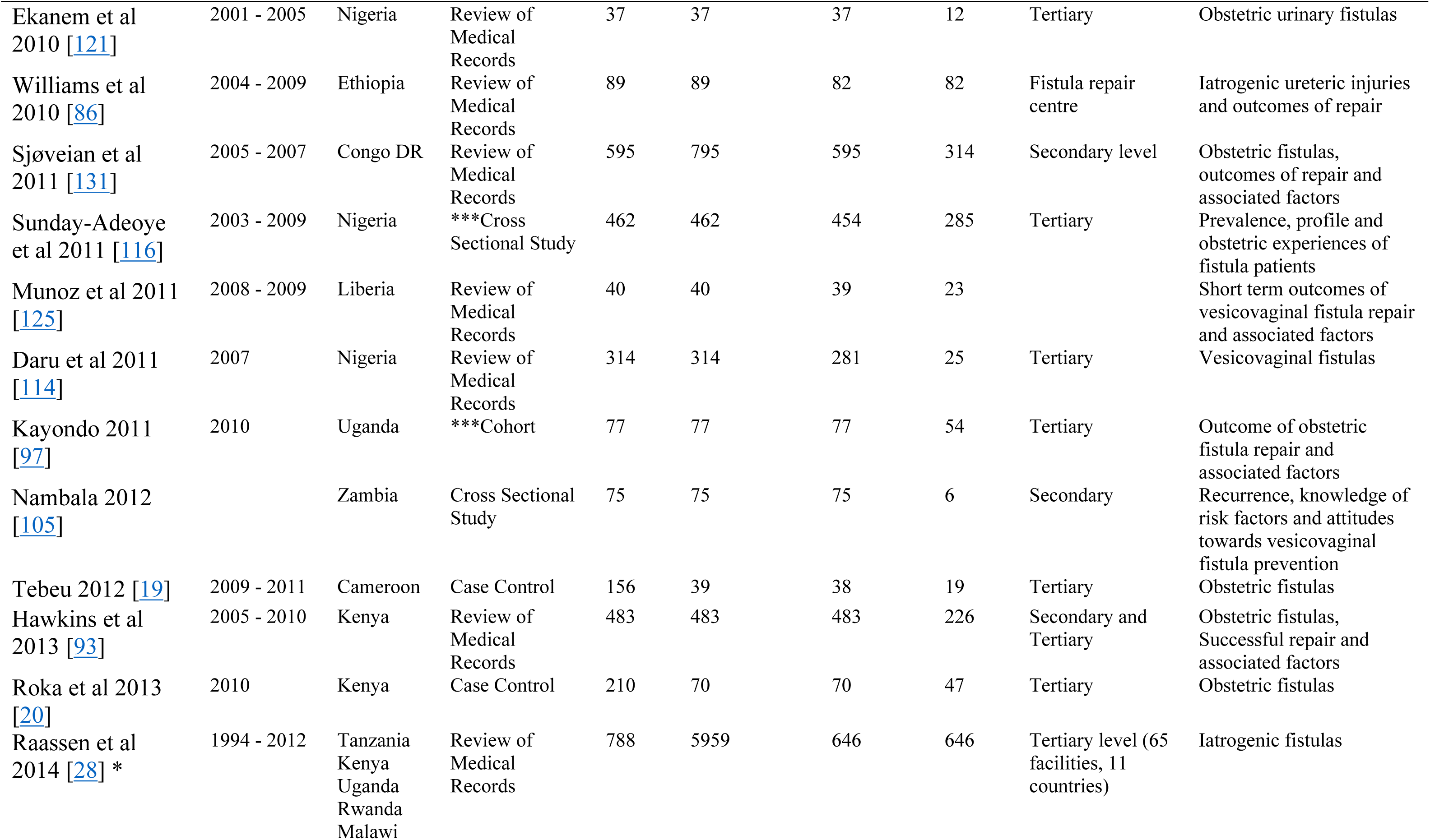
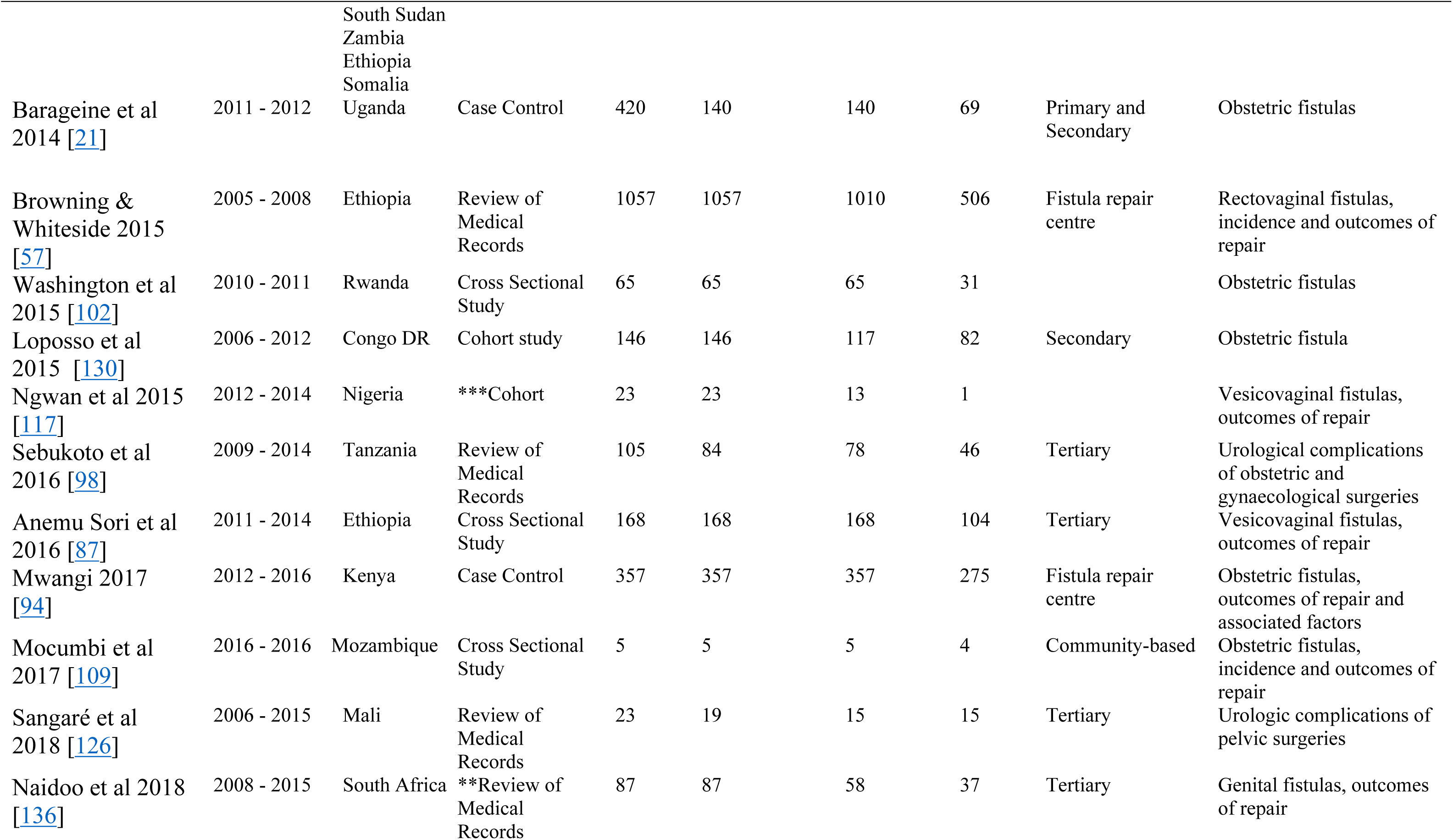
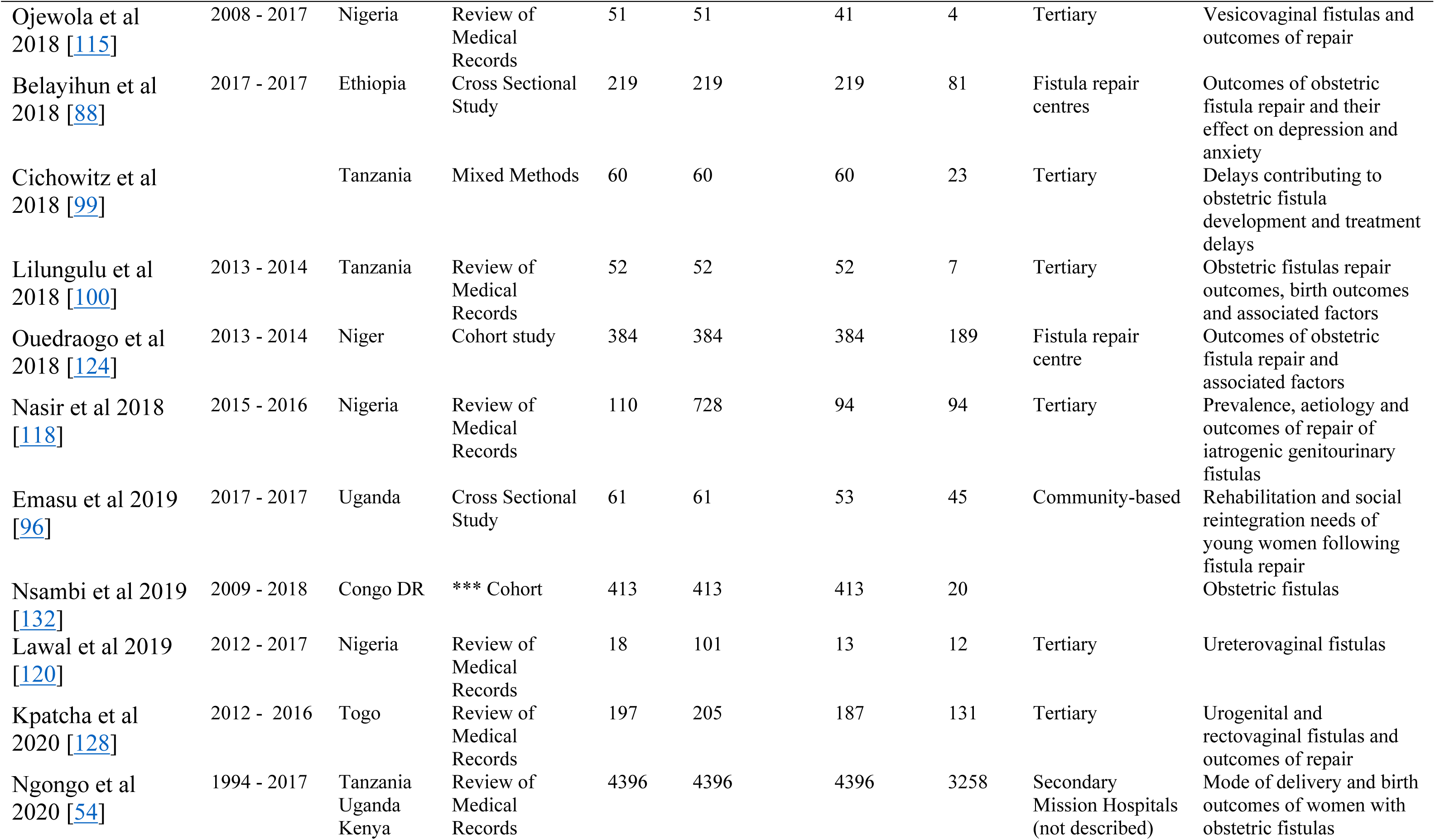
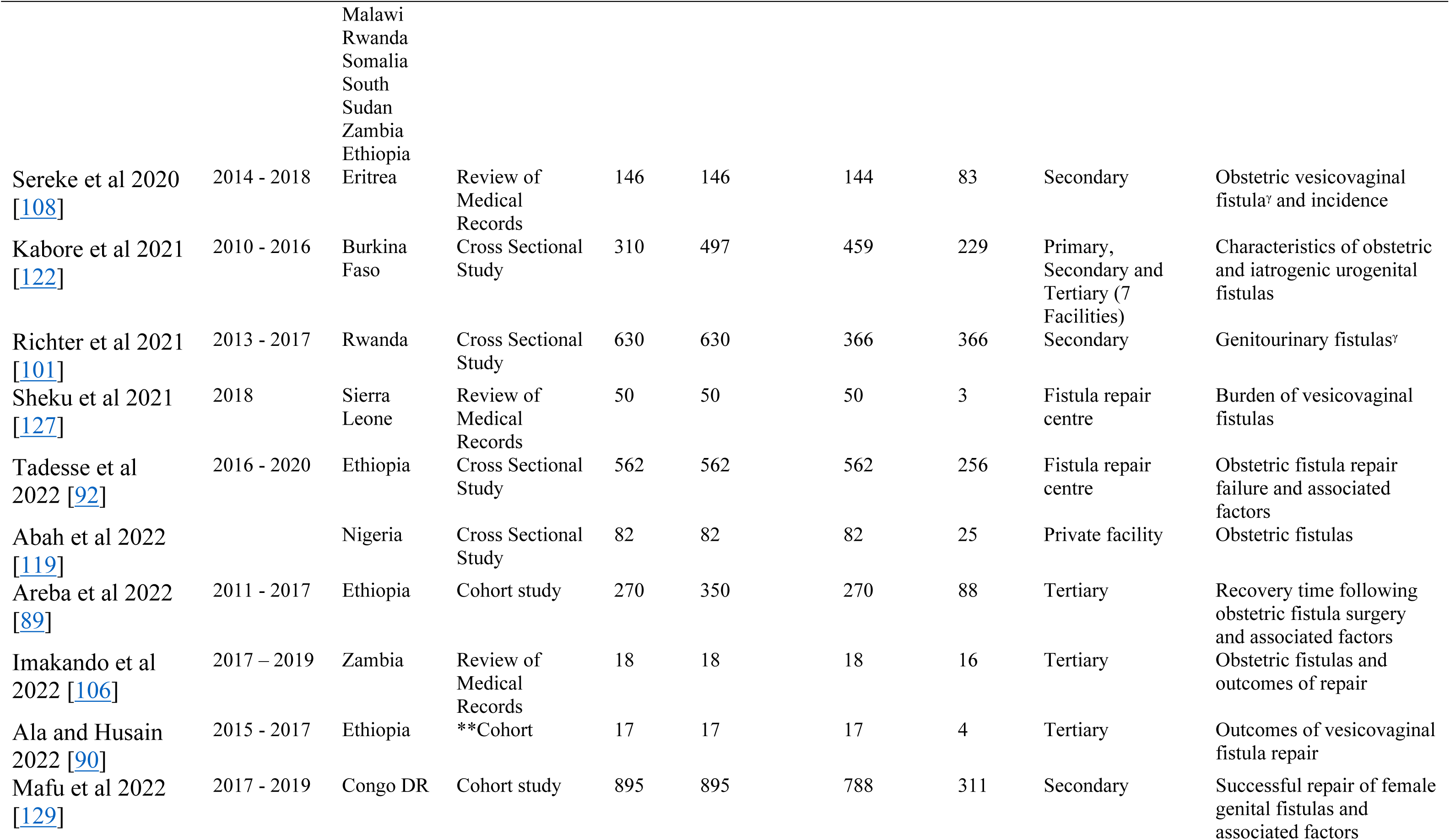
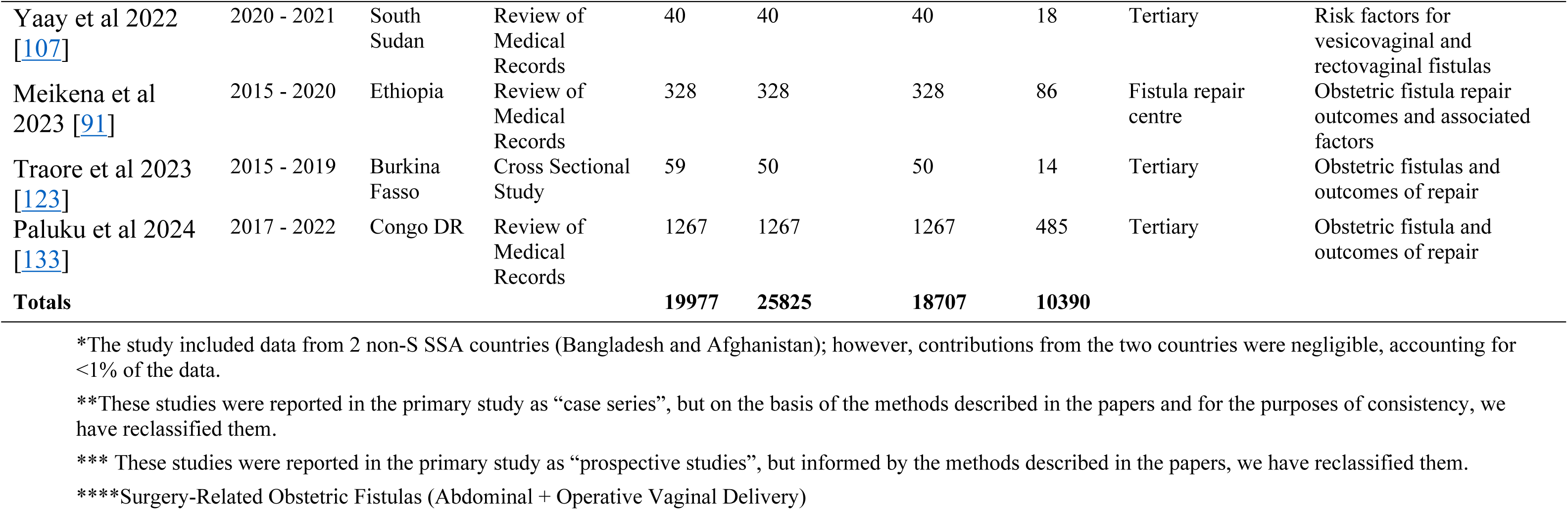
Characteristics of the included studies.

#### Study setting

The included studies were conducted in 22 sub-Saharan African countries across four regions, East, West, Central and South Africa, on the basis of the United Nations classification [49]. East Africa reported the greatest number of studies (30 studies): 8 studies from Ethiopia [57, 86–92], four from Kenya [20, 93–95], three from Uganda [21, 96, 97], three from Tanzania [98–100], three from Rwanda [101–103], three from Zambia [104–106], one from South Sudan [107], one from Eritrea [108] and one from Mozambique [109]. Three additional studies [28, 54, 110] were multicentre studies across several East African countries. Two of the multicentre studies [28, 54] involved nine East Africa countries, namely, Tanzania, Uganda, Kenya, Rwanda, Malawi, South Sudan, Zambia, Ethiopia and Somalia, whereas the other study [110] involved three countries, namely, Kenya, Tanzania and Uganda. Twenty-one studies were conducted in West Africa: 13 in Nigeria [59, 61, 111–121], two in Burkina Faso [122, 123], two in Niger [17, 124], one each in Liberia [125], Mali [126], Sierra Leone [127] and Togo [128]. Eight studies were conducted in Central Africa, five from the Democratic Republic of Congo [82, 129–133], two from Cameroon [22, 134] and one from the Republic of the Congo [135]. Two studies were from southern Africa, and both were conducted in South Africa [136, 137].

Over half of the studies were conducted at tertiary facilities (n=31) [19, 20, 59, 61, 87, 89, 90, 97–100, 103, 106, 107, 111–118, 120, 121, 123, 126, 128, 133, 135–137], eight studies at secondary facilities [22, 101, 104, 105, 108, 129–131], four studies at mixed primary and tertiary facilities [28, 93, 110, 122], one study at primary and secondary facilities [21], one study at primary facilities and mission hospitals (level of care unclear) [54] and nine studies [17, 57, 86, 88, 91, 92, 94, 124, 127] at dedicated fistula hospitals, whereas two were community-based studies [96, 109]. One study was conducted at a mission hospital (the level is unclear) [125], and one was conducted at a private facility [119]. For two studies, the study site was not indicated [102, 132].

#### Types of studies

All the studies were observational and were based on a review of medical records (30 studies) [28, 54, 57, 59, 61, 86, 91, 93, 98, 100, 104, 106–108, 111–115, 118, 120, 121, 125–128, 131, 133, 135, 136], cross-sectional studies (14 studies) [17, 22, 87, 88, 92, 96, 101, 102, 105, 109, 116, 119, 122, 123], cohort studies (11 studies) [89, 90, 97, 103, 110, 117, 124, 129, 130, 132, 137], case‒control studies (four studies) [20, 21, 94, 134] and mixed methods studies (one study) [99].

### Patient demographic and obstetric characteristics

The youngest person who developed an obstetric fistula was 11 years old [131], whereas the oldest patient was 66 years old [102]. Among the 13 studies reporting residences [14, 88, 89, 91, 100, 109, 122, 123, 127, 129, 133, 135, 137], 3217 women out of 4015 (80.1%) lived in rural areas, translating to 4 in 5 women with obstetric fistulas residing in rural areas. Twenty-two studies [20–22, 87, 89, 91–93, 100, 104, 109, 119, 121, 123, 124, 128, 131–135, 137] reporting on parity differentiated primiparous women from multiparous women. Among the 5507 obstetric fistulas, 2484 (45.1%) were prime-parous. Two studies [88, 107] reported 96 of the 259 obstetric fistulas (37.1%) and did not differentiate paras fistulas and two but grouped them together. Another study [90] grouped paras one, two and three together. Among the 14 studies reporting antenatal attendance [21, 22, 88, 89, 92, 97, 104, 105, 119, 121, 123, 127, 134, 137] of 1942 women with obstetric fistulas, 61.9% (n=1203) had attended antenatal clinics at a formal health facility at least once. A third of the women (33.7%) did not receive any form of antenatal care. Only 16.1% (n=194) had 4 or more antenatal visits.

Thirty-two studies [20, 22, 57, 61, 87–89, 91, 92, 94, 96, 97, 99, 102, 104, 105, 107–112, 118, 119, 123, 124, 127, 131–134, 137] reported the place of delivery leading to fistula development. Over two-thirds (68.4%) of the women delivered at a health facility, whereas 26.5% delivered outside a health facility. Among the non-health facility deliveries, 97.3% (n=2389) were home deliveries. Other deliveries occurred in transit to the health facility [57, 110]. Eight studies [21, 93, 104, 105, 107, 111, 121, 127] reported that attending at delivery preceded fistula development (n=1983). Skilled birth attendants were involved in 63.6% (n=1262) of deliveries, whereas 24.7% (n=490) were attended to by unskilled personnel. Most of these (98.2%) were traditional birth attractants (TBAs). The rest of the women were either delivered by relatives [105] or had no delivery attendant [21].

All but five studies [28, 86, 101, 118, 126] reported both vaginal and abdominal deliveries (n=17504). Among these, 56.3% (n=9846) were vaginal deliveries. Among the vaginal deliveries, 80.4% (n=7915) were spontaneous vaginal deliveries, whereas 18.9% (n=1858) were instrumental vaginal deliveries (either forceps or vacuum). Symphysiotomies and destructive vaginal procedures were conducted in <1% (n=72) of the vaginal deliveries. Operative deliveries were recorded in 41.9% (n=7329) of obstetric fistulas. Over 95.7% of the operative deliveries were Caesarean sections, 2.3% were Caesarean hysterectomies, and 1.8% had total or subtotal hysterectomies for various reasons, such as a ruptured uterus [103], intractable postpartum haemorrhage following vaginal delivery [133] and puerperal sepsis [137].

#### Birth outcomes

There were 33 studies reporting birth outcomes [17, 20–22, 28, 54, 57, 87, 88, 90–93, 96, 97, 99, 100, 102–104, 106–109, 119, 124, 127, 128, 130–133, 137], with 12158 women with obstetric fistulas. The still birth rate was 76.9% (n=9355), and the live birth rate was 17.8% (n=2170). Of these, seven studies [54, 99, 100, 102, 108, 128, 131] reported the outcomes for vaginal (n=2416) and caesarean sections (n=3019) separately. The live birth rate was higher (18.7%) among women who delivered by caesarean section than among those who delivered vaginally (11.9%).

### Meta-Analysis

#### Proportion of iatrogenic obstetric fistulas

The pooled proportion of iatrogenic obstetric fistulas among genitourinary and rectovaginal fistulas for sub-Saharan Africa was estimated to be 40% (95% CI 34%–47%, 60 studies, n=25825) (Fig 2), and the pooled proportion of iatrogenic obstetric fistulas among obstetric fistulas was 44% (95% CI 37–51%, 55 studies, n=17504) (Fig 3). There was considerable variation in the proportion of reported fistulas across studies, ranging from 2% to 89% for iatrogenic obstetric fistulas among genitourinary and rectovaginal fistulas and 2–92% for iatrogenic obstetric fistulas in women with obstetric fistulas.

**Fig 2.**
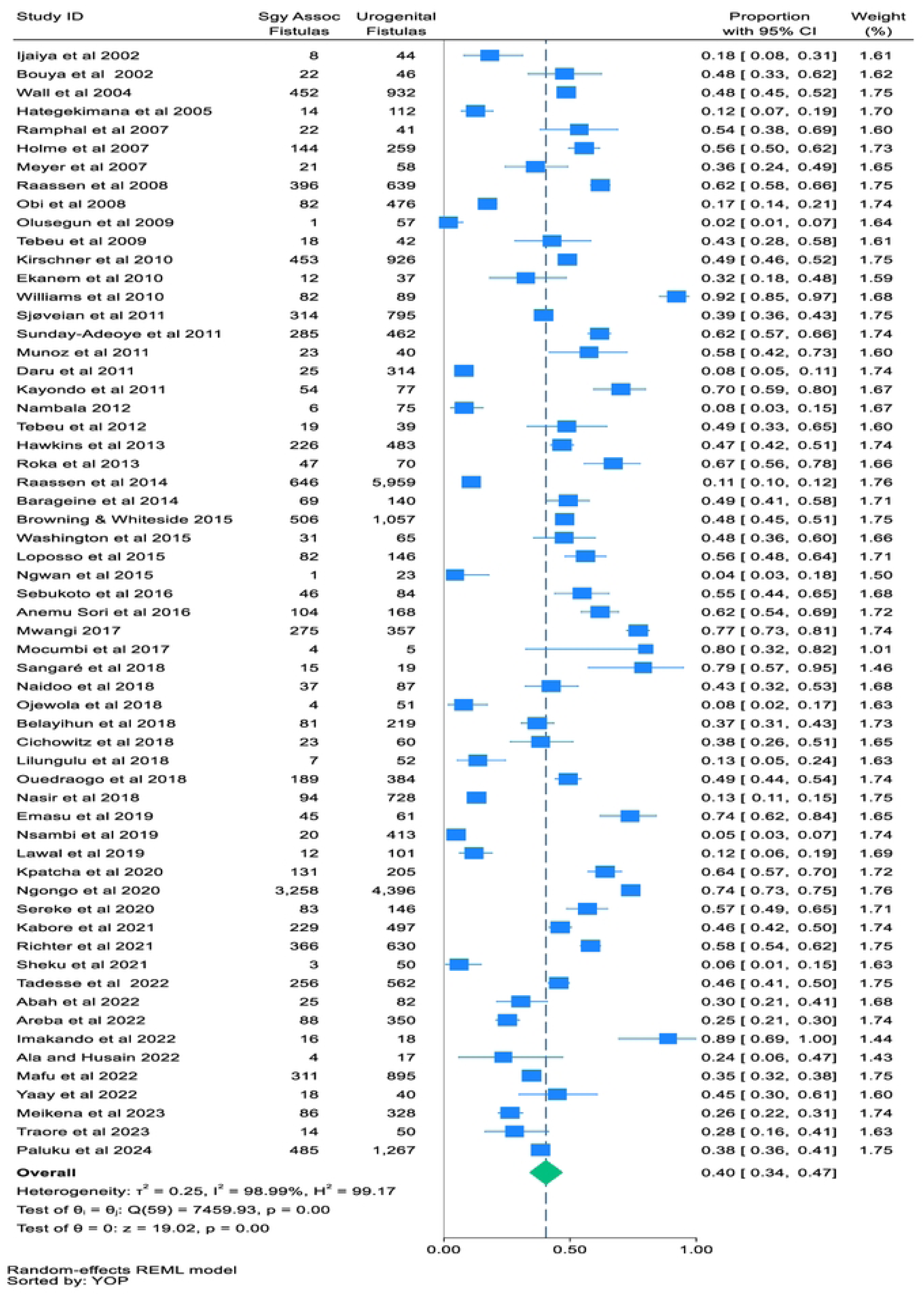
Proportion of iatrogenic obstetric fistulas among urogenital and rectovaginal fistulas.

**Fig 3.**
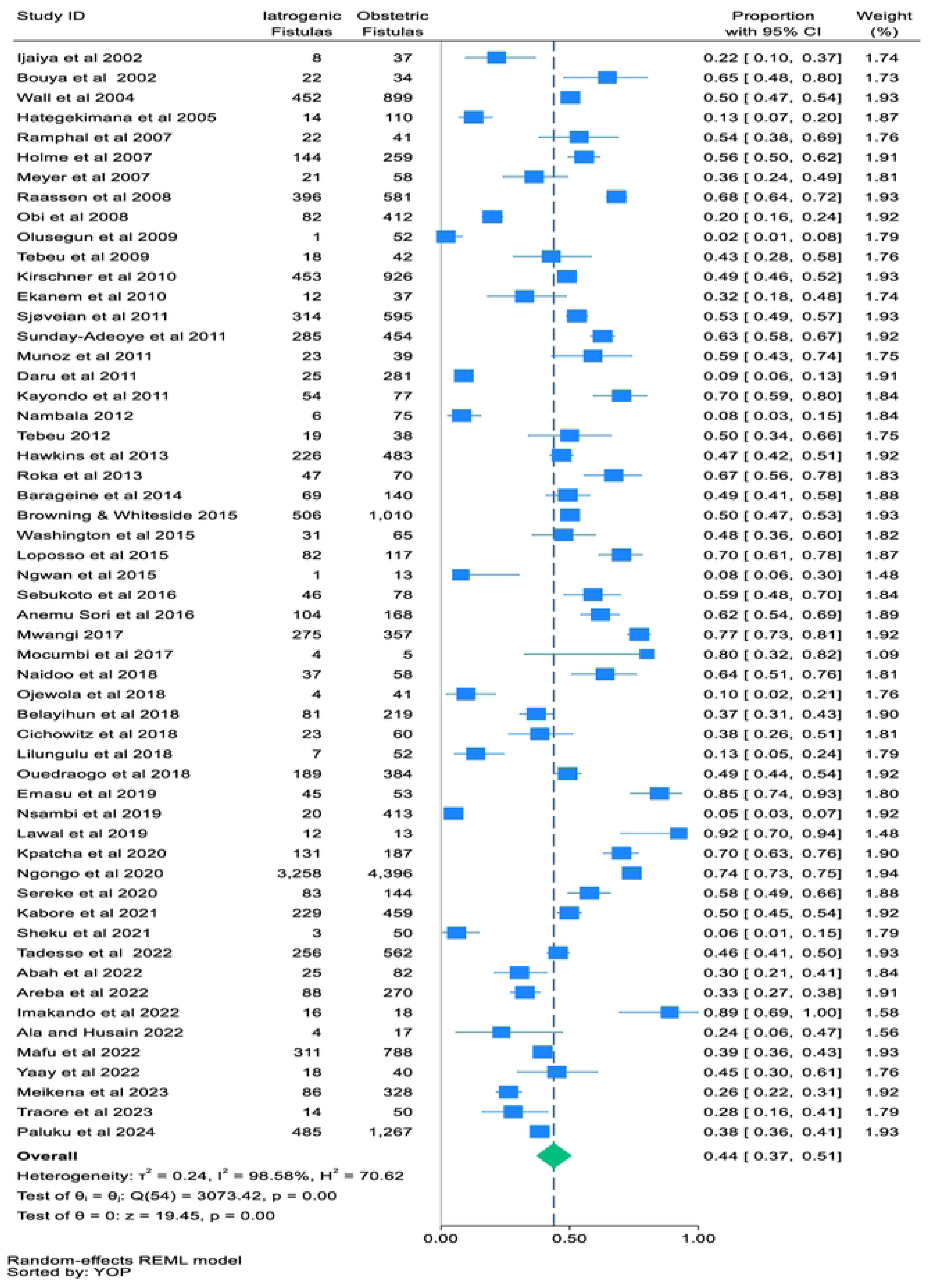
Proportion of iatrogenic obstetric fistulas among obstetric fistulas.

### Proportion of iatrogenic obstetric fistula by region of SSA

There was subregional variation in the proportion of surgery-related obstetric fistulas: East Africa at 48% (95% CI 39–58%; 26 studies, n=9572), Central Africa at 44% (95% CI 28–60%; eight studies, n=3294) and West Africa at 37% (95% CI 25–49%; 19 studies, n=4539) (Fig 4). Southern Africa has the highest proportion at 60% (95% CI 50–69%; two studies, n=99). The findings from southern Africa, however, came from only two small studies.

**Fig 4.**
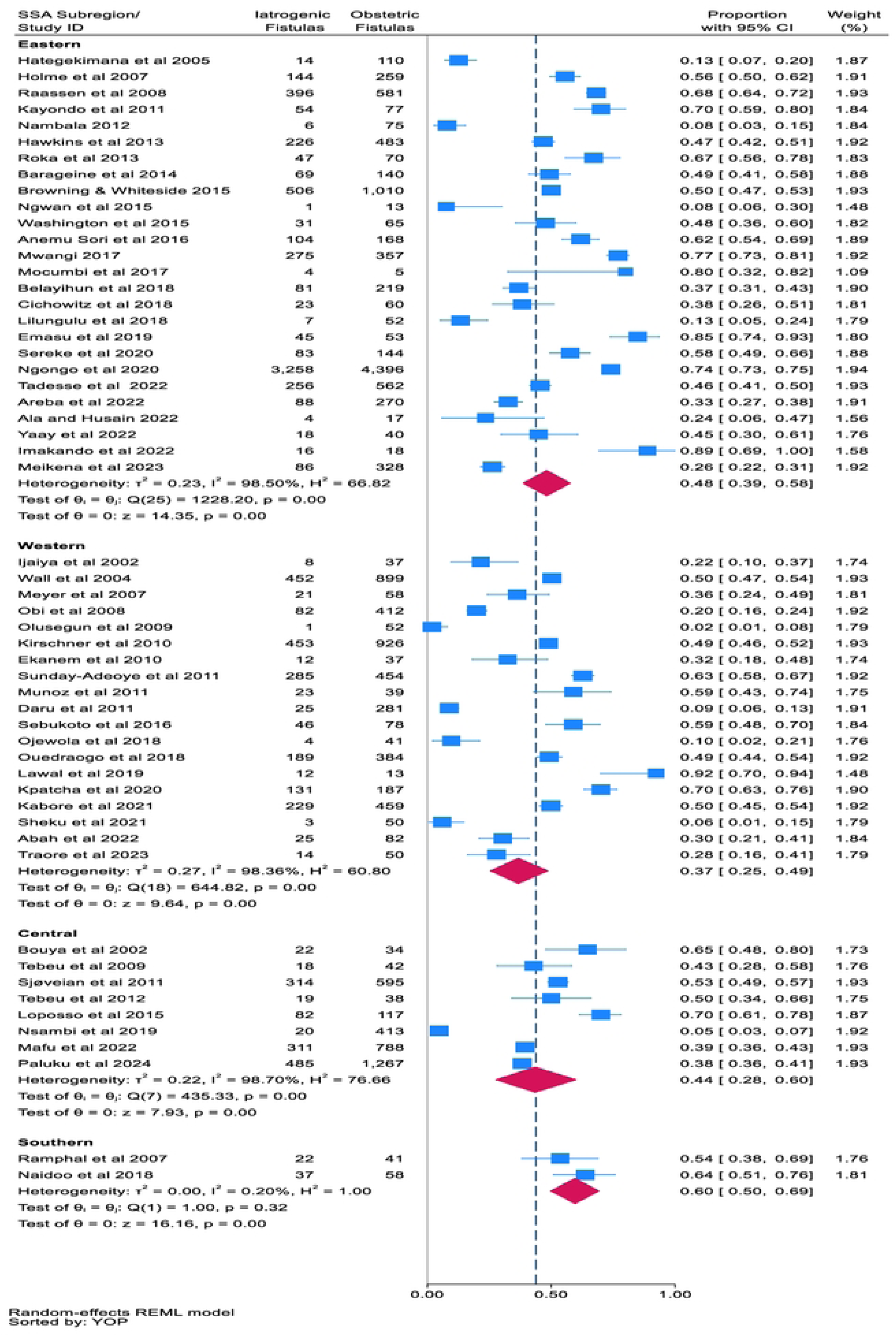
Proportion of iatrogenic obstetric fistulas by region of sub-Saharan Africa.

### Proportion of iatrogenic obstetric fistulas by year of publication of the study

There is no demonstrable trend in the proportion of obstetric fistulas over the past two decades (2002–2024), with proportions averaging approximately 40% (Fig 5).

**Fig 5.**
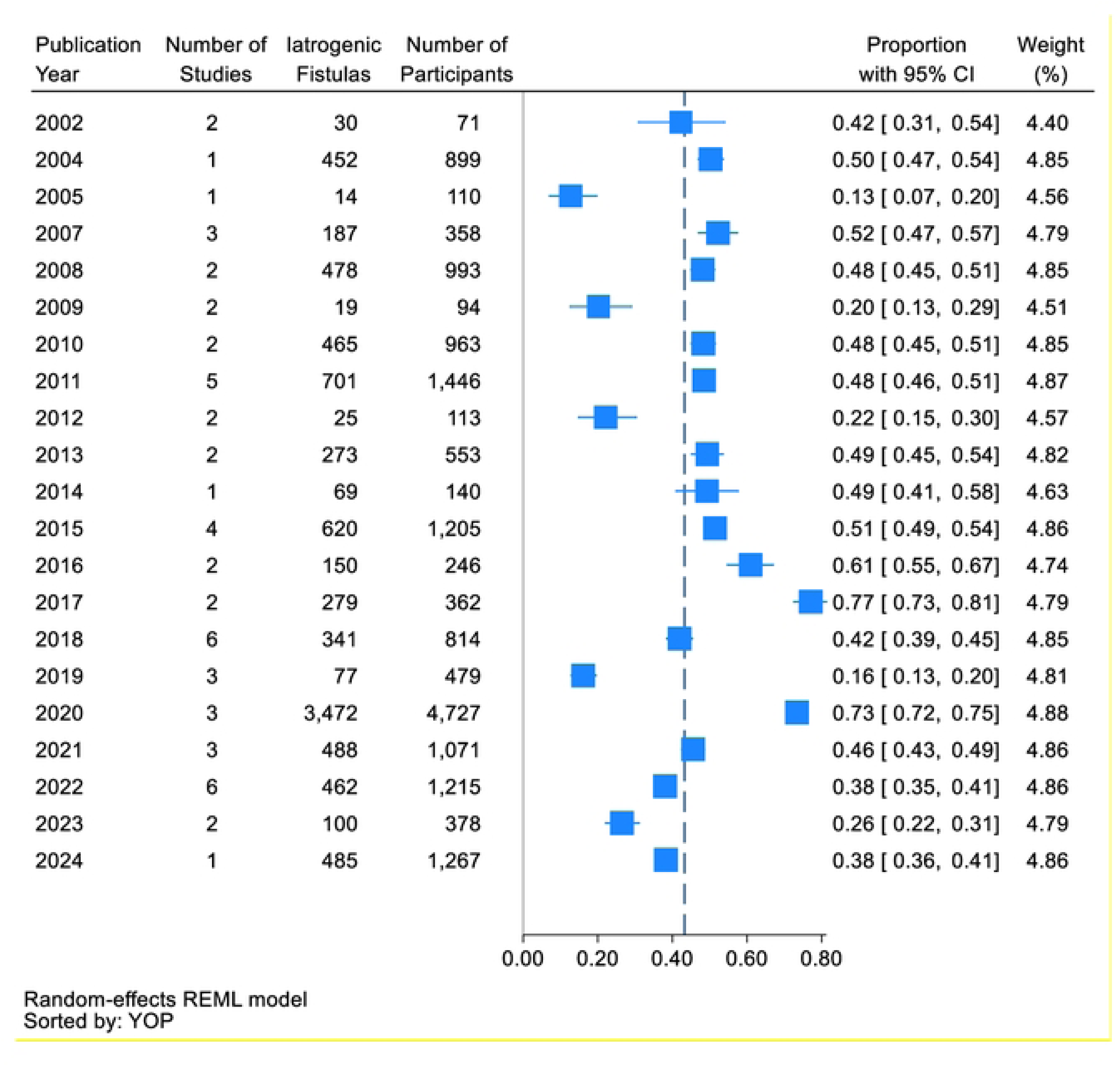
Proportion of iatrogenic obstetric fistulas among women with obstetric fistulas by publication year.

### Subgroup by type of iatrogenic fistula

The types of fistulas are reported in Fig 6a (with an extension in Fig 6b). Using the classification by Raassen et al. [28], the proportion of obstetric fistulas that are definitely iatrogenic (ureteric/ureterovaginal) was found to be 7% (95% CI 3–12%, 13 studies, n=4273), whereas the proportion of obstetric fistulas that are probably (vault) iatrogenic was estimated to be 13% (95% CI 11–16%, one study, n=646) [28]. The proportion of vesicouterine fistulas was estimated to be 14% (95% CI 5–26%, 10 studies, n=3864) (Fig 6a). There are insufficient data to differentiate between likely iatrogenic and likely iatrogenic vesicouterine fistulas, as birth outcomes are not classified by fistula type. Fistulas that are not included in the classification by Raassen et al. [28] are outlined in Fig 6b. The proportion of vesicovaginal fistulas was estimated to be 47% (95% CI 37–57%, 30 studies, n=7726), whereas the proportion of rectovaginal fistulas was estimated to be 36% (95% CI 25–48%, 13 studies, n=4639). The incidence of the combination of vesicovaginal and rectovaginal fistulas was estimated to be 43% (95% CI 32–54%, 10 studies, n=4051), whereas that of urethrovaginal fistulas was estimated to be 40% (95% CI 33–48%, five studies, n=1803).

**Fig 6a.**
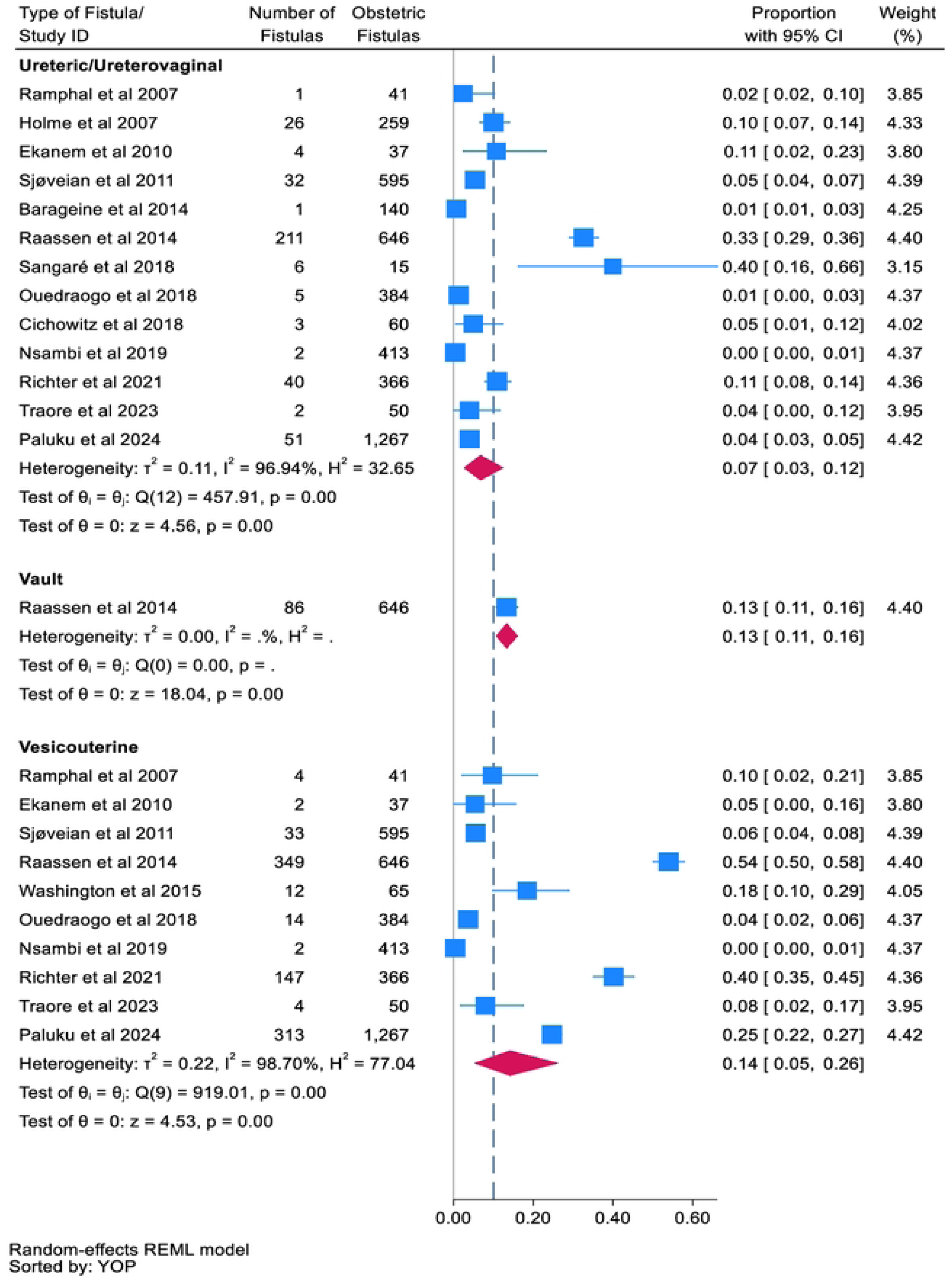
Subgroup analysis by fistula type.

**Fig 6b.**
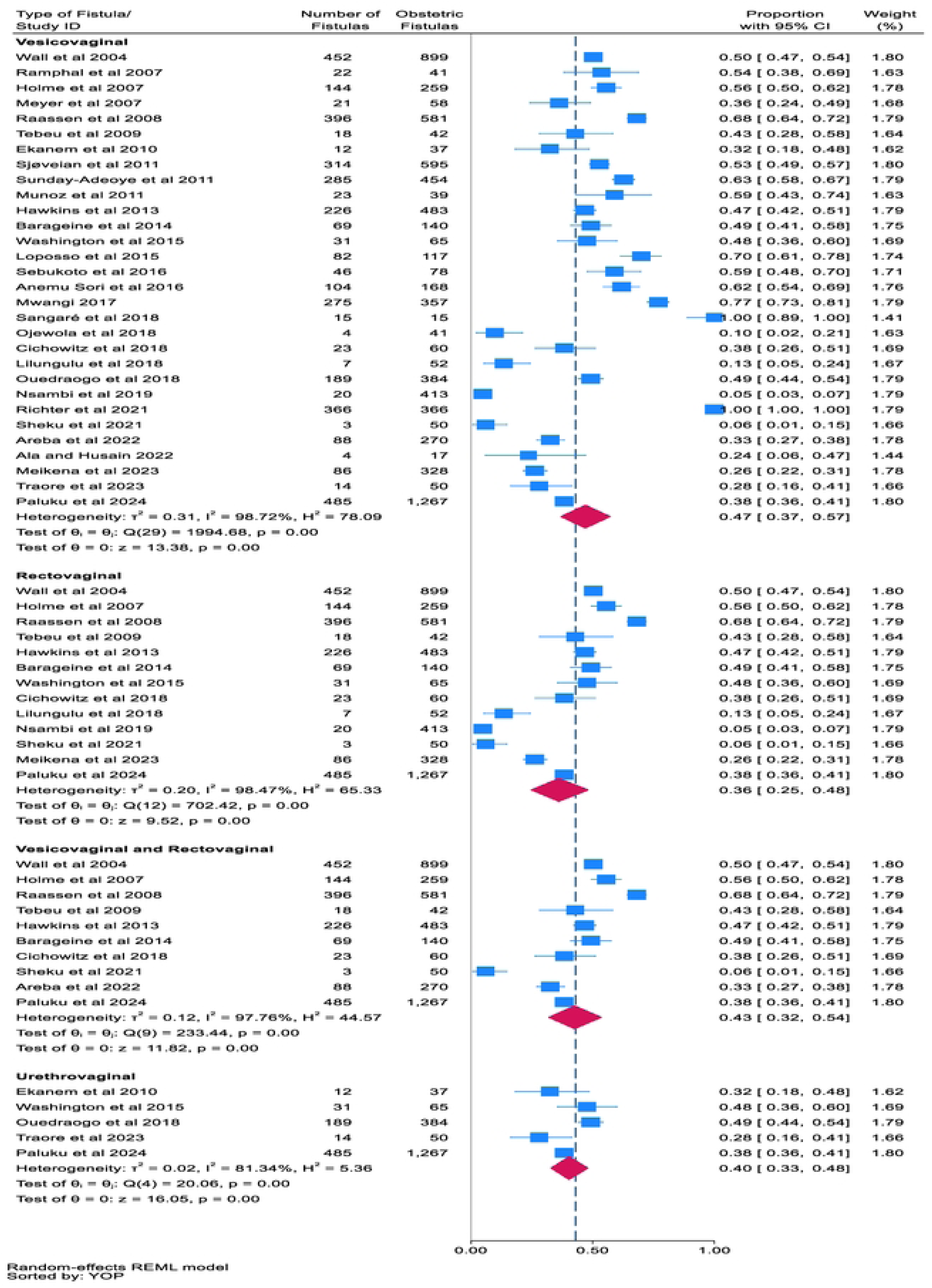
Subgroup analysis by fistula type.

### Types of obstetric surgery preceding fistulas

The types of obstetric surgery preceding obstetric fistulas are reported in Fig 7a (with an extension from Fig 7b). Caesarean section was the most common obstetric procedure preceding obstetric fistula, accounting for 34% (95% CI 28–40%, 54 studies, n=17491) (Fig 7a), followed by instrumental vaginal delivery 11% (95% CI 9–13%, 34 studies, n=12727), caesarean hysterectomy 8% (95% CI 2–17%, 7 studies, n=1988), total/subtotal hysterectomy 7% (95% CI 4–10%; five studies, n=1533), and repair of ruptured uterus 6% (95% CI 2–12%, 5 studies, n=355) (Fig 7b).

**Fig 7a.**
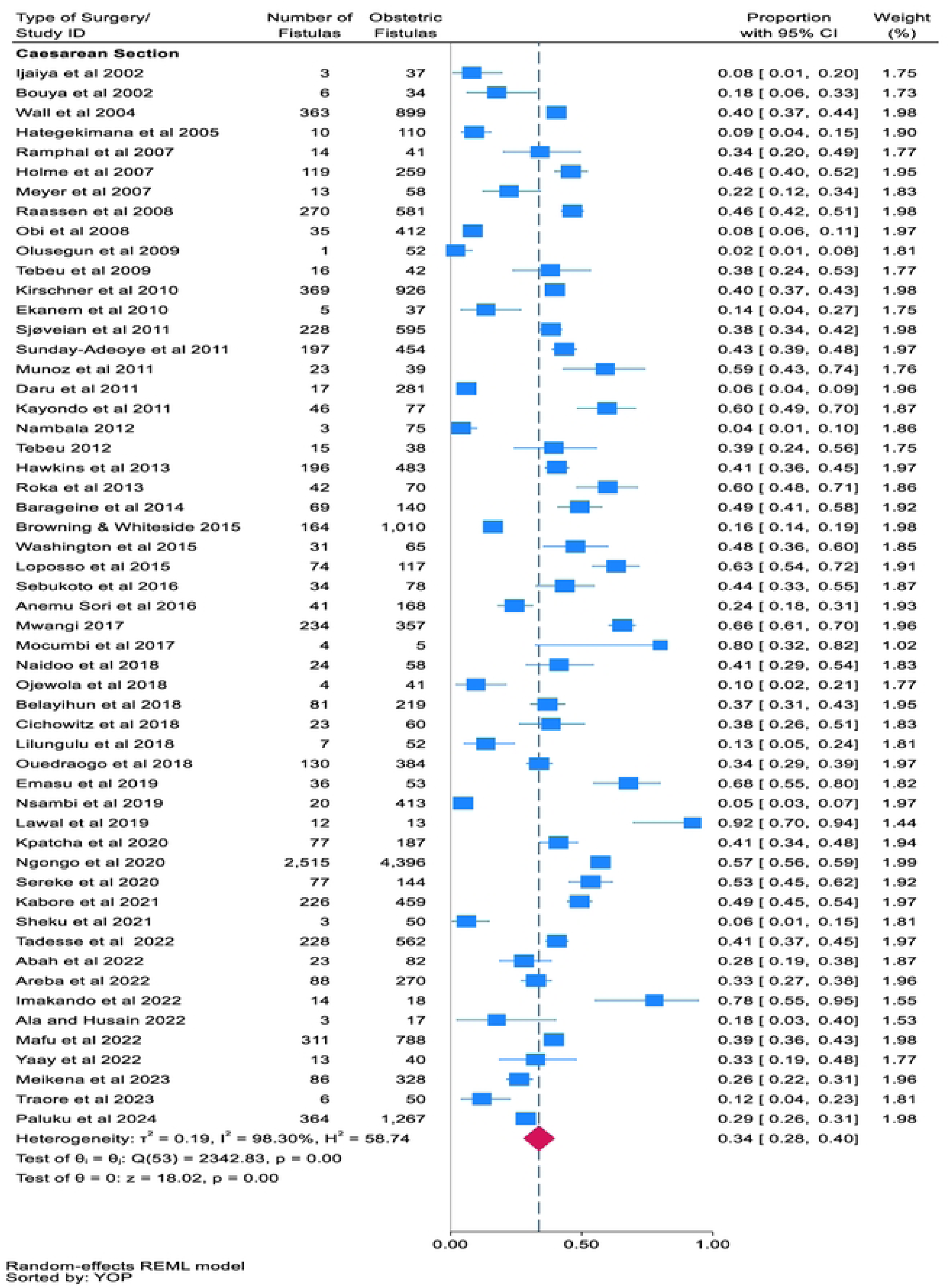
Type of surgery preceding fistulas.

**Fig 7b.**
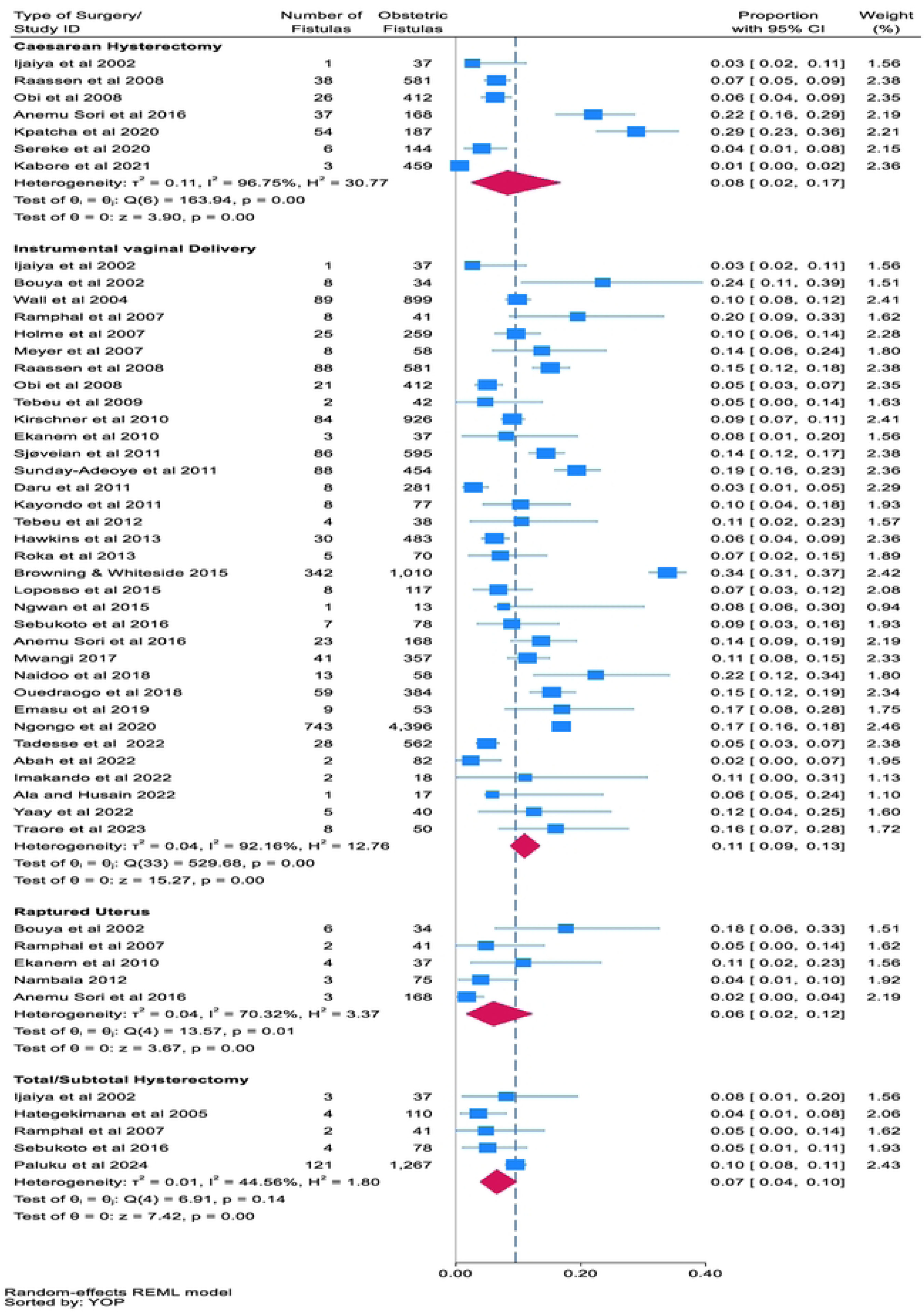
Type of surgery preceding fistulas.

### Personnel performing surgeries

Four studies [21, 28, 122, 136] reported the type of personnel who conducted surgeries preceding fistulas. One study [122] was excluded from the analysis, as the personnel conducting the surgeries were reported collectively for both obstetric and non-obstetric surgeries. Among the remaining studies, nonspecialists accounted for 92% (95% CI 61–100%, three studies, n=752), and specialists accounted for 3% (95% 1–5%, two studies, n=683) (Fig 8).

**Fig 8.**
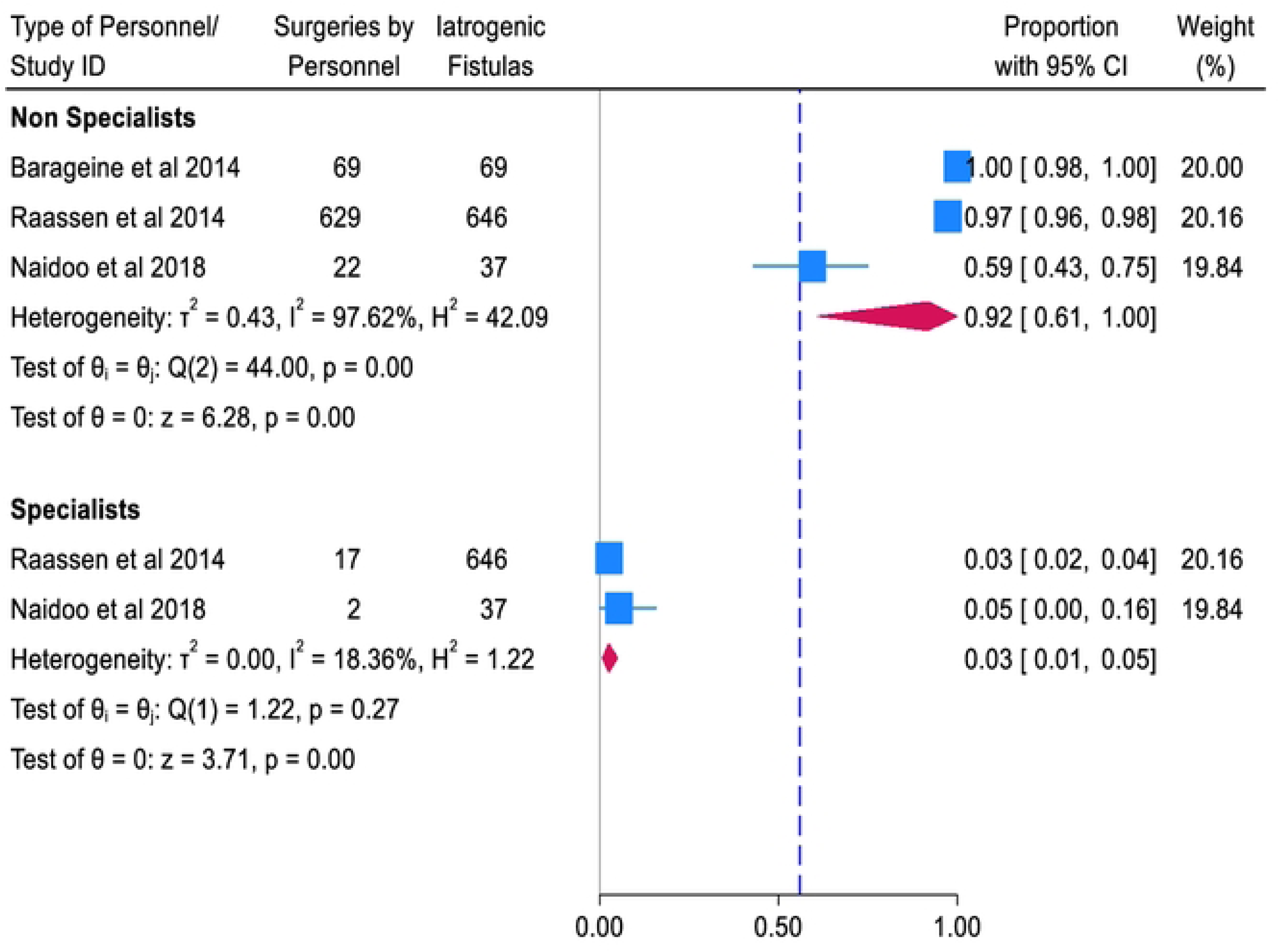
Types of personnel conducting surgery preceding fistulas.

Subgroup analysis by rural urban distribution could not be conducted because the data on the iatrogenic fistulas were not stratified by rural urban area.

### Risk of bias results

The results of the risk of bias in the included studies are reported in Table 2. With respect to the study’s target population as a representative sample of the national population, only five studies [20, 21, 94, 109, 134] were judged as having a low risk of bias. The remaining 55 studies were judged as having a high risk of bias. Given that the sampling frame is a true or close representation of the target population, all the studies were judged as having a low risk of bias except one [96], whose risk of bias was unclear. With respect to random sampling or a census being undertaken, two studies were judged as having an unclear risk of bias, whereas the remaining 58 studies were judged as having a low risk of bias. All the studies were judged as having a low risk of bias for the likelihood of nonresponse bias, whether the data were collected directly from subjects (or a proxy was used) and whether an acceptable case definition was used in the study. With respect to the validity and reliability of the study instrument that was used to measure the parameter of interest, seven studies [88, 91, 92, 96, 105, 116, 121] were judged as having a low risk of bias. The remaining studies were judged as having an unclear risk of bias. With respect to the use of the same mode of data collection for all study participants, the appropriateness of the length of the shortest proportion period for the parameter of interest and the appropriateness of the numerator(s) and denominators, all studies were judged to have a low risk of bias. Overall, all the studies were rated as having a low risk of bias (see Table 2).

**Table 2.**
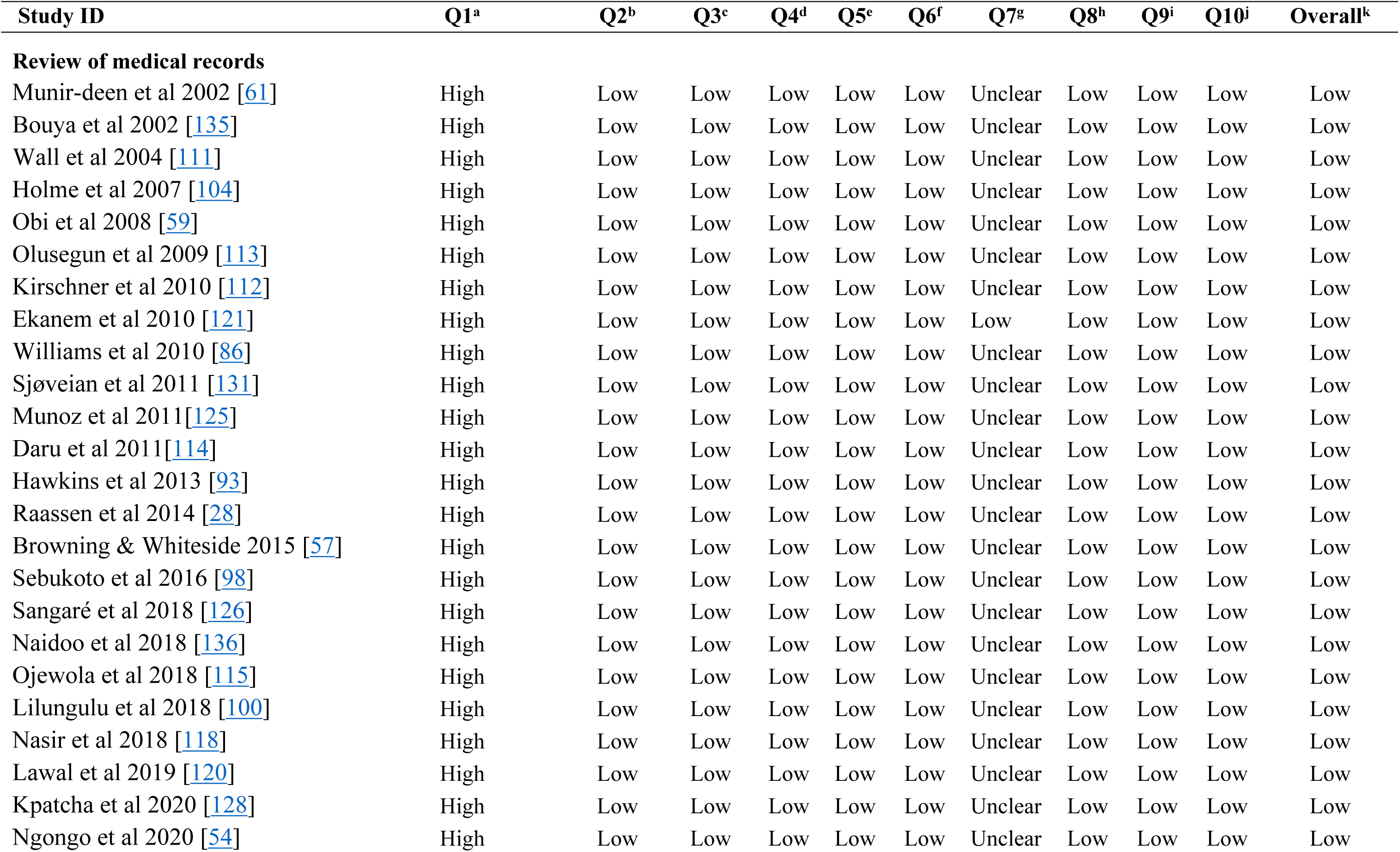
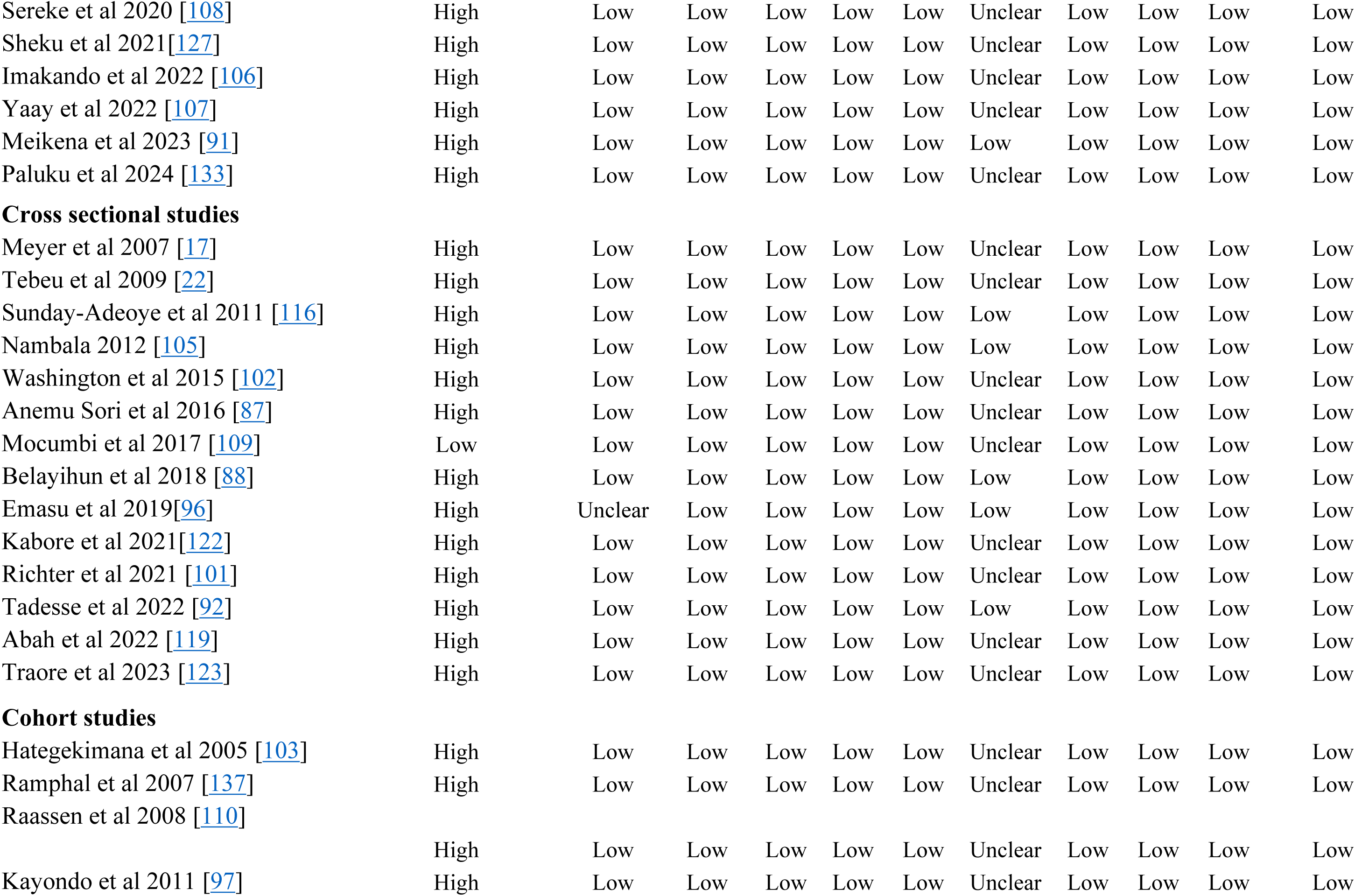
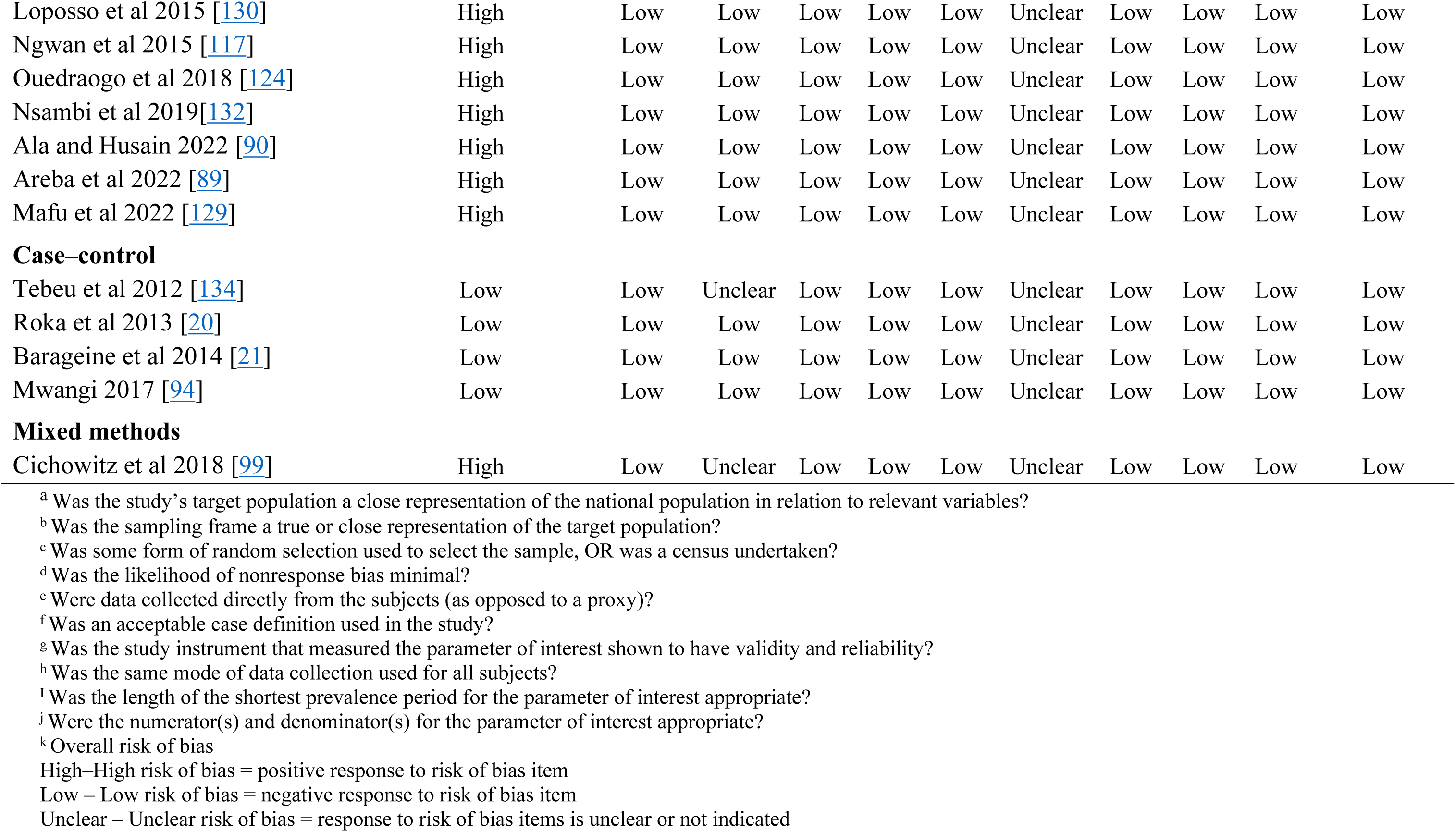
Risk of bias in the included studies.

| Study ID | Q1<sup>a</sup> | Q2<sup>b</sup> | Q3<sup>c</sup> | Q4<sup>d</sup> | Q5<sup>e</sup> | Q6<sup>f</sup> | Q7<sup>g</sup> | Q8<sup>h</sup> | Q9<sup>i</sup> | Q10<sup>j</sup> | Overall<sup>k</sup> |
| --- | --- | --- | --- | --- | --- | --- | --- | --- | --- | --- | --- |
| **Review of medical records** |  |  |  |  |  |  |  |  |  |  |  |
| Munir-deen et al 2002 [61] | High | Low | Low | Low | Low | Low | Unclear | Low | Low | Low | Low |
| Bouya et al 2002 [135] | High | Low | Low | Low | Low | Low | Unclear | Low | Low | Low | Low |
| Wall et al 2004 [111] | High | Low | Low | Low | Low | Low | Unclear | Low | Low | Low | Low |
| Holme et al 2007 [104] | High | Low | Low | Low | Low | Low | Unclear | Low | Low | Low | Low |
| Obi et al 2008 [59] | High | Low | Low | Low | Low | Low | Unclear | Low | Low | Low | Low |
| Olusegun et al 2009 [113] | High | Low | Low | Low | Low | Low | Unclear | Low | Low | Low | Low |
| Kirschner et al 2010 [112] | High | Low | Low | Low | Low | Low | Unclear | Low | Low | Low | Low |
| Ekanem et al 2010 [121] | High | Low | Low | Low | Low | Low | Low | Low | Low | Low | Low |
| Williams et al 2010 [86] | High | Low | Low | Low | Low | Low | Unclear | Low | Low | Low | Low |
| Sjøveian et al 2011 [131] | High | Low | Low | Low | Low | Low | Unclear | Low | Low | Low | Low |
| Munoz et al 2011 [125] | High | Low | Low | Low | Low | Low | Unclear | Low | Low | Low | Low |
| Daru et al 2011 [114] | High | Low | Low | Low | Low | Low | Unclear | Low | Low | Low | Low |
| Hawkins et al 2013 [93] | High | Low | Low | Low | Low | Low | Unclear | Low | Low | Low | Low |
| Raassen et al 2014 [28] | High | Low | Low | Low | Low | Low | Unclear | Low | Low | Low | Low |
| Browning & Whiteside 2015 [57] | High | Low | Low | Low | Low | Low | Unclear | Low | Low | Low | Low |
| Sebukoto et al 2016 [98] | High | Low | Low | Low | Low | Low | Unclear | Low | Low | Low | Low |
| Sangaré et al 2018 [126] | High | Low | Low | Low | Low | Low | Unclear | Low | Low | Low | Low |
| Naidoo et al 2018 [136] | High | Low | Low | Low | Low | Low | Unclear | Low | Low | Low | Low |
| Ojewola et al 2018 [115] | High | Low | Low | Low | Low | Low | Unclear | Low | Low | Low | Low |
| Lilungulu et al 2018 [100] | High | Low | Low | Low | Low | Low | Unclear | Low | Low | Low | Low |
| Nasir et al 2018 [118] | High | Low | Low | Low | Low | Low | Unclear | Low | Low | Low | Low |
| Lawal et al 2019 [120] | High | Low | Low | Low | Low | Low | Unclear | Low | Low | Low | Low |
| Kpatcha et al 2020 [128] | High | Low | Low | Low | Low | Low | Unclear | Low | Low | Low | Low |
| Ngongo et al 2020 [54] | High | Low | Low | Low | Low | Low | Unclear | Low | Low | Low | Low |
| Sereke et al 2020 [108] | High | Low | Low | Low | Low | Low | Unclear | Low | Low | Low | Low |
| Sheku et al 2021 [127] | High | Low | Low | Low | Low | Low | Unclear | Low | Low | Low | Low |
| Imakando et al 2022 [106] | High | Low | Low | Low | Low | Low | Unclear | Low | Low | Low | Low |
| Yaay et al 2022 [107] | High | Low | Low | Low | Low | Low | Unclear | Low | Low | Low | Low |
| Meikena et al 2023 [91] | High | Low | Low | Low | Low | Low | Low | Low | Low | Low | Low |
| Paluku et al 2024 [133] | High | Low | Low | Low | Low | Low | Unclear | Low | Low | Low | Low |
| **Cross sectional studies** |  |  |  |  |  |  |  |  |  |  |  |
| Meyer et al 2007 [17] | High | Low | Low | Low | Low | Low | Unclear | Low | Low | Low | Low |
| Tebeu et al 2009 [22] | High | Low | Low | Low | Low | Low | Unclear | Low | Low | Low | Low |
| Sunday-Adeoye et al 2011 [116] | High | Low | Low | Low | Low | Low | Low | Low | Low | Low | Low |
| Nambala 2012 [105] | High | Low | Low | Low | Low | Low | Low | Low | Low | Low | Low |
| Washington et al 2015 [102] | High | Low | Low | Low | Low | Low | Unclear | Low | Low | Low | Low |
| Anemu Sori et al 2016 [87] | High | Low | Low | Low | Low | Low | Unclear | Low | Low | Low | Low |
| Mocumbi et al 2017 [109] | Low | Low | Low | Low | Low | Low | Unclear | Low | Low | Low | Low |
| Belayihun et al 2018 [88] | High | Low | Low | Low | Low | Low | Low | Low | Low | Low | Low |
| Emasu et al 2019 [96] | High | Unclear | Low | Low | Low | Low | Low | Low | Low | Low | Low |
| Kabore et al 2021 [122] | High | Low | Low | Low | Low | Low | Unclear | Low | Low | Low | Low |
| Richter et al 2021 [101] | High | Low | Low | Low | Low | Low | Unclear | Low | Low | Low | Low |
| Tadesse et al 2022 [92] | High | Low | Low | Low | Low | Low | Low | Low | Low | Low | Low |
| Abah et al 2022 [119] | High | Low | Low | Low | Low | Low | Unclear | Low | Low | Low | Low |
| Traore et al 2023 [123] | High | Low | Low | Low | Low | Low | Unclear | Low | Low | Low | Low |
| **Cohort studies** |  |  |  |  |  |  |  |  |  |  |  |
| Hategekimana et al 2005 [103] | High | Low | Low | Low | Low | Low | Unclear | Low | Low | Low | Low |
| Ramphal et al 2007 [137] | High | Low | Low | Low | Low | Low | Unclear | Low | Low | Low | Low |
| Raassen et al 2008 [110] | High | Low | Low | Low | Low | Low | Unclear | Low | Low | Low | Low |
| Kayondo et al 2011 [97] | High | Low | Low | Low | Low | Low | Unclear | Low | Low | Low | Low |
| Loposso et al 2015 [130] | High | Low | Low | Low | Low | Low | Unclear | Low | Low | Low | Low |
| Ngwan et al 2015 [117] | High | Low | Low | Low | Low | Low | Unclear | Low | Low | Low | Low |
| Ouedraogo et al 2018 [124] | High | Low | Low | Low | Low | Low | Unclear | Low | Low | Low | Low |
| Nsambi et al 2019[132] | High | Low | Low | Low | Low | Low | Unclear | Low | Low | Low | Low |
| Ala and Husain 2022 [90] | High | Low | Low | Low | Low | Low | Unclear | Low | Low | Low | Low |
| Areba et al 2022 [89] | High | Low | Low | Low | Low | Low | Unclear | Low | Low | Low | Low |
| Mafu et al 2022 [129] | High | Low | Low | Low | Low | Low | Unclear | Low | Low | Low | Low |
| Case-control |  |  |  |  |  |  |  |  |  |  |  |
| Tebeu et al 2012 [134] | Low | Low | Unclear | Low | Low | Low | Unclear | Low | Low | Low | Low |
| Roka et al 2013 [20] | Low | Low | Low | Low | Low | Low | Unclear | Low | Low | Low | Low |
| Barageine et al 2014 [21] | Low | Low | Low | Low | Low | Low | Unclear | Low | Low | Low | Low |
| Mwangi 2017 [94] | Low | Low | Low | Low | Low | Low | Unclear | Low | Low | Low | Low |
| Mixed methods |  |  |  |  |  |  |  |  |  |  |  |
| Cichowitz et al 2018 [99] | High | Low | Unclear | Low | Low | Low | Unclear | Low | Low | Low | Low |

<sup>a</sup> Was the study's target population a close representation of the national population in relation to relevant variables?
<sup>b</sup> Was the sampling frame a true or close representation of the target population?
<sup>c</sup> Was some form of random selection used to select the sample, OR was a census undertaken?
<sup>d</sup> Was the likelihood of nonresponse bias minimal?
<sup>e</sup> Were data collected directly from the subjects (as opposed to a proxy)?
<sup>f</sup> Was an acceptable case definition used in the study?
<sup>g</sup> Was the study instrument that measured the parameter of interest shown to have validity and reliability?
<sup>h</sup> Was the same mode of data collection used for all subjects?
<sup>i</sup> Was the length of the shortest prevalence period for the parameter of interest appropriate?
<sup>j</sup> Were the numerator(s) and denominator(s) for the parameter of interest appropriate?
<sup>k</sup> Overall risk of bias
High-High risk of bias = positive response to risk of bias item
Low – Low risk of bias = negative response to risk of bias item
Unclear – Unclear risk of bias = response to risk of bias items is unclear or not indicated

## Discussion

### Summary of the main results

The majority of women who develop obstetric fistulas in SSA are rural residents (4 in 5 women) with suboptimal antenatal care. Nearly half (45%) of these women are primiparous at the time of fistula development. Approximately 40% of urogenital/rectovaginal fistulas and 44% of obstetric fistulas are associated with obstetric surgeries. The burden of surgery-related obstetric fistulas somewhat varies across regions, with Western Africa having the lowest proportion at 37% and Central Africa and Eastern Africa having similar proportions at 44% and 48%, respectively. Southern Africa has the highest proportion at 60%; however, caution should be taken in interpreting this finding, as it was based on only two small studies conducted in one country (South Africa). The most common obstetric surgeries preceding fistula development were caesarean sections (34%), followed by instrumental vaginal deliveries (11%). Hysterectomy following caesarean section preceded 8% of obstetric fistulas, whereas total/subtotal hysterectomy preceded 7% of cases. Six percent of women with obstetric fistulas successfully repaired the ruptured uterus. Among fistula types, vesicouterine fistulas are the most common at 14%, followed by vault fistulas at 13% and ureteric fistulas at 7%. The proportion of vault fistulas is, however, based on a single study. Notably, 92% of obstetric surgeries preceding fistula development were conducted by nonspecialists, raising concerns about the quality of surgical care in the region.

### Completeness and applicability of the evidence

The high proportion of iatrogenic obstetric fistulas in the region is consistent with the observed increases in the rates of caesarean delivery [26], with high rates of surgical task shifting/sharing to meet the demand [32, 138]. The global rise in caesarean deliveries varies widely, with lower rates, 8.2%, in the least developed countries, the majority of which are in SSA, compared with 24.2% in less developed countries and 27.2% in more developed countries [139]. Despite higher caesarean section rates, obstetric fistulas are rare in developed countries [140, 141]. The critical difference may be attributed to task sharing, a common phenomenon in developing countries. Arguments in favour of task sharing include lower costs of training and deployment and higher rates of retention in rural areas [142]. However, there is a need to ensure that the gains made by task sharing [143] are not negated by complications akin to reduced surgical quality.

The results of the present review indicate that more than 9 of the 10 surgeries preceding fistula development are performed by nonspecialists in SSA. This suggests inequities in access to specialist obstetric care services and the critical shortage of specialists (obstetricians and gynecologists) in the region. Obstetric fistulas are more common in patients referred from district facilities, which are largely staffed by medical officers with limited obstetric experience [136]. Often, medical officers conducting surgeries have had as little as 3 months of internship training in an obstetric/gynaecology unit [21, 144, 145]. The limited time left gaps in obstetric surgical skill acquisition, affecting the quality of surgical care [146].

The inequity gap is further widened by rural urban disparities. Four of the five women with obstetric fistulas were noted to be rural residents in the present review. Inequities in access to maternal health services in SSA disadvantage rural populations [147] as well as the urban poor [148]. Critical shortages in human resources, long distances to health facilities, financial constraints and cultural influences limit access to skilled birth attendants in underserved rural communities [149]. Within-country variations in access to emergency surgical procedures could explain intra-country variations in the prevalence of surgery-associated obstetric fistulas. In Nigeria, this percentage ranges from 2% [117] to 92% [120], whereas in Ethiopia, it ranges from 24% [90] to 62% [87]. Congo DR ranged from 5% [132] to 70% [130]. This may be due to within-country variations in access to emergency obstetric surgical procedures. These inequity gaps need to be bridged to achieve universal maternal health coverage [150]. At the subregional level, further inquiry is needed into the situation in southern Africa, as only one country, South Africa, was represented, with low power due to its small sample size.

The proportion of surgery-related fistulas among obstetric fistulas ranged from 13% in 2004 to 77% in 2017. The information obtained from these studies was inadequate for revealing an association between the incidence of iatrogenic obstetric fistulas and the year of publication. However, despite the variations by year, the findings indicate that iatrogenic obstetric fistulas have been consistently present in the past two decades. This raises the need to address this public health issue, without which a substantial caseload of fistulas will remain for years to come, even if fistulas from prolonged obstructed labour are eliminated [16].

The majority of iatrogenic obstetric fistulas are associated with caesarean delivery, with or without hysterectomy, and instrumental vaginal delivery. The fistula subtypes reported in the present study are consistent with the surgical aetiology. Ureteric fistulas, which are definitely iatrogenic, constitute 7%, whereas vault fistulas, which are probably iatrogenic, constitute 13% of obstetric fistulas. Vesicouterine fistulas, which can be either probable or likely iatrogenic depending on whether the birth outcome is good or poor, constitute 14% of obstetric fistulas.

There is an urgent need to scale up programs aimed at increasing the number of obstetricians/gynaecologists to mitigate the critical shortage in SSA, particularly in rural and remote areas. Additional interventions should be aimed at improving the quality of obstetric surgical care through targeted training programs at the undergraduate level and in service [151]. Considerations for training programs should include the development of a consensus on the “minimum acceptable obstetric training time” to inform quality assurance standards for clinicians involved in the provision of surgical obstetric care or develop/upscale programs for continued surgical skill acquisition at facilities with no resident obstetricians. Skills should include competence-based training in operative vaginal deliveries.

### Quality of the evidence

The quality of the included studies was assessed via the tool developed by Hoy et al. With the exception of one study whose data were based on a subset of a population-based survey [109] and four case‒control studies [20, 21, 94, 134], the studies were not representative of population data, as the majority were based on participants presenting to health facilities and/or facility-based data. However, this is unlikely to have affected the quality of the estimates on the proportion of iatrogenic obstetric fistulas among both the urogenital/rectovaginal fistulas and obstetric fistulas, since, owing to the nature of obstetric fistulas, patients are generally hard to come by and therefore facility-based data are a good proxy for the national population. There was a low risk of selection bias, as nearly all the studies involved total enumeration sampling or census. Additionally, all the studies utilized data collected directly from participants or medical records of participants themselves with appropriate case definitions for the study outcomes, and data collection among individual studies was uniform. Four studies [88, 91, 96, 105] used validated data collection instruments, whereas three studies [92, 116, 121] used instruments that were pretested. For the remaining studies, it was unclear whether the instruments used for data collection were validated or pretested. This could be due to the retrospective nature of the majority of studies. However, despite these variations, the study findings were consistent with sociodemographic, obstetric and fistula characteristics and were therefore unlikely to affect the quality of outcomes. Overall, all the studies were judged as having a low risk of bias for the outcomes of interest.

### Potential biases in the review process

This systematic review followed standard guidelines and best practices, and used comprehensive and rigorous methods published in a protocol that sought to minimize all forms of biases encountered during the systematic review process. During data extraction, two studies [17, 103] reported data as percentages from the sample size, without absolute figures, posing a minor challenge in data extraction; however, this is unlikely to have introduced major bias in the study.

### Agreements with existing evidence

The proportion of surgical procedures preceding iatrogenic fistulas in SSA is similar to the findings of a study conducted in Asia, which attributed 37.6% to caesarean sections and 7.6% to instrumental deliveries [30]. In an assessment of iatrogenic urogenital fistulas of gynaecological origin in a low-income setting outside SSA, 89% of surgeries were performed by nonspecialists [152]. This highlights challenges in the quality of both obstetric and gynaecological surgical care in developing countries.

Previous studies have compared outcomes of caesarean section between physicians and nonphysicians (such as clinical officers and medical licentiates) and noted no significant differences between the two groups [142, 153]. More recently, a study noted no significant increase in the risk of iatrogenic obstetric fistulas between physicians and associated clinicians [154]. However, data comparing the risk for fistula development and other caesarean-related complications between nonspecialists (associating clinicians and general medical practitioners) and specialists in the region before the present study were scarce.

### Strengths and limitations of the overall evidence

Whereas most studies have compared the risk for fistula development and other caesarean-related complications between physicians and nonphysicians, to the best of our knowledge, this is the first study to compare nonspecialists (associate clinicians and general medical practitioners) with specialists in relation to fistula development. This review has several limitations resulting from quality of the studies included as most them were based on facility data and not surveys. However, since this was uniform across nearly all studies, it is unlikely that this would have affected the magnitude and direction of the pooled (overall) estimates. The review findings did not differentiate obstetric procedures after prolonged labour from those occurring without labour (elective procedures), as the majority of the included studies did make such a differentiation. Despite this limitation, the ureteric, vault and vesicouterine fistulas are consistent with iatrogenic injury as opposed to ischaemic injury from prolonged obstructed labour. An additional limitation was the absence of data on the years of surgical experience for the different types of personnel was not available and this could not be accounted for in the meta-analysis.

## Conclusion

The available evidence indicates a high proportion of surgery-related obstetric fistulas among urogenital/rectovaginal fistulas and obstetric fistulas in sub-Saharan Africa (40% and 44%, respectively). The most common surgery type antecedent to fistulas is caesarean section, followed by instrumental vaginal delivery, over 92% of which are performed by nonspecialists. The fact that more than 90% of obstetric surgeries are performed by nonspecialists and that the incidence of iatrogenic fistulas is high calls for increased efforts towards capacity building and quality improvement across countries in SSA. Interventions should be tailored to improve access to quality obstetric surgical care for high-risk subpopulations, namely, rural residents and women in their prime pregnancies. Further research that combines the type of personnel and level of surgical experience is needed to determine the best practices for performing different obstetric operations, i.e., total or subtotal hysterectomies, caesarean sections in women with previous caesarean deliveries and caesarean sections in women with prolonged obstructed labour, whose tissues are more susceptible to iatrogenic injury.

## Data Availability

All relevant data are within the manuscript and its Supporting Information files.

## List of abbreviations

CI: Confidence interval
CS: Caesarean Section
EmOC: Emergency obstetric care
LMICs: Low- and middle-income countries
PRISMA: Preferred Reporting Items for Systematic Reviews and Meta-analyses
PROSPERO: International Prospective Register for Systematic Reviews
RCT: Randomized Controlled Trial
SDGs: Sustainable development goals
SSA: Sub-Saharan Africa
TBA: traditional birth attractant
UN: United Nations
UNFPA: United Nations Population Fund
VCVF: Vesicervical-Vaginal Fistulas
WHO: World Health Organization

## Additional Files

None

## Declarations

### Ethics approval and consent to participate

The study did not require ethical clearance, as it involved the use of secondary data.

### Consent for publication

Not applicable

### Availability of data and materials

All the data generated or analysed during this study are included in this published article.

## Acknowledgements

This systematic review was prepared as part of the capacity-building initiative of the Centre for Evidence Synthesis and Policy (CESP), University of Ghana and the Africa Communities of Evidence Synthesis and Translation (ACEST), which train health professionals and scientists in evidence synthesis and translation across countries in Africa and low- and middle-income countries (LMICs). Dr. Mercy Monde Imakando is an obstetrician gynaecologist specializing in Evidence Synthesis and Translation; she is mentored by Prof Anthony Danso-Appiah (Director, Centre for Evidence Synthesis and Policy, University of Ghana).

## Author Contributions

Conceptualization: MMI, EM, ADA

Investigation: MMI, EM, DO, MMW, ADA

Methodology: MMI, EM, DO, MMW, CJ, KOA, IF, ADA

Resources: MMI, EM, DO, MMW, CJ, KOA, IF, ADA

Validation: MMI, EM, DO, MMW, CJ, KOA, IF, ADA

Writing - original draft: MMI, EM, DO, MMW, CJ, KOA, ADA

Writing - review and editing of manuscript: MMI, EM, DO, MMW, CJ, KOA, IF, ADA

Supervision: ADA

## REFERENCES

1. UNFPA. Obstetric Fistula: United Nations Population Fund 2022 [updated 23 May 2022. Available from: https://www.unfpa.org/obstetric-fistula.

2. Polan ML, Sleemi A, Bedane MM, Lozo S, Morgan MA. Obstetric Fistula. In: Debas HT, Donkor P, Gawande A, Jamison DT, Kruk ME, Mock CN, editors. Essential Surgery: Disease Control Priorities, Third Edition (Volume 1). Washington (DC): The International Bank for Reconstruction and Development / The World Bank; 2015.

3. WHO. Obstetric Fistula: Worl Health Organisation; 2018 [updated 2018 February 19 cited 2021 July 6]. Available from: https://www.who.int/news-room/facts-in-pictures/detail/10-facts-on-obstetric-fistula.

4. Miller S, Lester F, Webster M, Cowan B. Obstetric fistula: a preventable tragedy. J Midwifery Womens Health 2005;50(4):286–94.

5. UNGA. UN Report on Obstetric Fistula 2020. UNFPA Campaign to End Fistula Website Inited Nations General Assembly; 2020 2020, July 28th Report No.: A/75/264 Contract No.: July 14th

6. Ahmed S, Tunçalp Ö. Burden of obstetric fistula: from measurement to action. The Lancet Global Health. 2015;3(5):e243–e4.

7. Geleto A, Chojenta C, Musa A, Loxton D. Barriers to access and utilization of emergency obstetric care at health facilities in sub-Saharan Africa: a systematic review of literature. Journal of Systematic Reviews. 2018;7(1):1–14.

8. Capes T, Ascher-Walsh C, Abdoulaye I, Brodman M. Obstetric fistula in low and middle income countries. Mount Sinai Journal of Medicine: A Journal of Translational Personalized Medicine. 2011;78(3):352–61.

9. Melah GS, Massa AA, Yahaya UR, Bukar M, Kizaya DD, El-Nafaty AU. Risk factors for obstetric fistulae in north-eastern Nigeria. Journal of obstetrics and gynaecology : the journal of the Institute of Obstetrics and Gynaecology. 2007;27(8):819–23.

10. Kelly J, Kwast B. Epidemiologic study of vesico-vaginal fistula in Ethiopia. International urogynecology journal. 1993;4:278–81.

11. Arrowsmith S, Hamlin EC, Wall LL. Obstructed labor injury complex: obstetric fistula formation and the multifaceted morbidity of maternal birth trauma in the developing world. Obstetrical & gynecological survey. 1996;51(9):568–74.

12. Creanga AA, Ahmed S, Genadry RR, Stanton C. Prevention and treatment of obstetric fistula: Identifying research needs and public health priorities. International Journal of Gynecology and Obstetrics. 2007;99:S151–S4.

13. Dangal G, Thapa K, Yangzom K, Karki A. Obstetric Fistula in the Developing World: An Agonising Tragedy. Nepal J Obstet Gynaecol. 2014;8.

14. Sih AM, Kopp DM, Tang JH, Rosenberg NE, Chipungu E, Harfouche M, et al. Association between parity and fistula location in women with obstetric fistula: a multivariate regression analysis. Bjog. 2016;123(5):831–6.

15. Reisenauer C. Presentation and management of rectovaginal fistulas after delivery. International urogynecology journal. 2016;27(6):859–64.

16. Fistula Care Plus. Iatrogenic Fistula: an Urgent Quality of Care Challenge: Fistula Care Plus, USAID, Engender Health; 2016 [Available from: https://fistulacare.org/wp-content/uploads/2015/10/Iatrogenic-fistula-technical-brief_2016-1.pdf.

17. Meyer L, Ascher-Walsh CJ, Norman R, Idrissa A, Herbert H, Kimso O, et al. Commonalities among women who experienced vesicovaginal fistulae as a result of obstetric trauma in Niger: results from a survey given at the National Hospital Fistula Center, Niamey, Niger. 2007;197(1):90. e1-. e4.

18. Wall LL. A framework for analyzing the determinants of obstetric fistula formation. Studies in Family Planning. 2012;43(4):255–72.

19. Tebeu PM, Fomulu JN, Khaddaj S, de Bernis L, Delvaux T, Rochat CH. Risk factors for obstetric fistula: a clinical review. International urogynecology journal. 2012;23(4):387–94.

20. Roka ZG, Akech M, Wanzala P, Omolo J, Gitta S, Waiswa P. Factors associated with obstetric fistulae occurrence among patients attending selected hospitals in Kenya, 2010: a case control study. BMC pregnancy and childbirth. 2013;13:56.

21. Barageine JK, Tumwesigye NM, Byamugisha JK, Almroth L, Faxelid E. Risk factors for obstetric fistula in Western Uganda: a case control study. PLoS One. 2014;9(11):e112299.

22. Tebeu PM, de Bernis L, Doh AS, Rochat CH, Delvaux T. Risk factors for obstetric fistula in the Far North Province of Cameroon. International journal of gynaecology and obstetrics: the official organ of the International Federation of Gynaecology and Obstetrics. 2009;107(1):12–5.

23. McCurdie FK, Moffatt J, Jones K. Vesicovaginal fistula in Uganda. Journal of obstetrics and gynaecology : the journal of the Institute of Obstetrics and Gynaecology. 2018;38(6):822–7.

24. Pope R. Research in obstetric fistula: addressing gaps and unmet needs. 2018;131(5):863–70.

25. Changole J, Thorsen VC, Kafulafula U. A road to obstetric fistula in Malawi: capturing women’s perspectives through a framework of three delays. Int J Womens Health. 2018;10:699–713.

26. Betrán AP, Ye J, Moller AB, Zhang J, Gülmezoglu AM, Torloni MR. The Increasing Trend in Caesarean Section Rates: Global, Regional and National Estimates: 1990-2014. PLoS One. 2016;11(2):e0148343.

27. El-Lamie IK. Urogenital fistulae: changing trends and personal experience of 46 cases. Int Urogynecol J Pelvic Floor Dysfunct. 2008;19(2):267–72.

28. Raassen TJ, Ngongo CJ, Mahendeka MM. Iatrogenic genitourinary fistula: an 18-year retrospective review of 805 injuries. International Urogynecology Journal. 2014;25(12):1699–706.

29. Onsrud M, Sjøveian S, Mukwege D. Cesarean delivery-related fistulae in the Democratic Republic of Congo. International journal of gynaecology and obstetrics: the official organ of the International Federation of Gynaecology and Obstetrics. 2011;114(1):10–4.

30. Tasnim N, Bangash K, Amin O, Luqman S, Hina H. Rising trends in iatrogenic urogenital fistula: A new challenge. International Journal of Gynecology and Obstetrics. 2020;148(S1):33–6.

31. Schneeberger C, Mathai M. Emergency obstetric care: Making the impossible possible through task shifting. International journal of gynaecology and obstetrics: the official organ of the International Federation of Gynaecology and Obstetrics. 2015;131 Suppl 1:S6–S9.

32. Federspiel F, Mukhopadhyay S, Milsom P, Scott JW, Riesel JN, Meara JG. Global surgical and anaesthetic task shifting: a systematic literature review and survey. Lancet (London, England). 2015;385 Suppl 2:S46.

33. Hoyler M, Hagander L, Gillies R, Riviello R, Chu K, Bergström S, Meara JG. Surgical care by non-surgeons in low-income and middle-income countries: a systematic review. Lancet (London, England). 2015;385 Suppl 2:S42.

34. WorldBank. Specialist Surgical Workforce (per 100, 000 population) worldbank.org: World Bank; 2021 [updated 2021 August 11. Available from: https://data.worldbank.org/indicator/SH.MED.SAOP.P5?end=2018&start=2010.

35. Kyei-Nimakoh M, Carolan-Olah M, McCann TV. Access barriers to obstetric care at health facilities in sub-Saharan Africa-a systematic review. Systematic reviews. 2017;6(1):110.

36. Imakando MM, Maya E, Owiredu D, Monde MW, Jacobs C, Fwemba I, et al. The burden of iatrogenic obstetric fistulas in Sub-Saharan Africa: Systematic review and meta-analysis protocol. PLoS One. 2024;19(8):e0302529.

37. Higgins JPT, Thomas J, Chandler J, Cumpston M, Li T, Page MJ, Welch VA. Cochrane Handbook for Systematic Reviews of Interventions: Cochrane; 2023.

38. Danso-Appiah A, Eusebi P, Lo N, Orso M, Akuffo KO, Fleming F, et al. Optimal Prevalence Threshold For Guiding The Implementation Of Preventive Chemotherapy In Countries Endemic For Schistosomiasis: Synthesis Of Evidence From Mass Drug Administration Programmes For Developing This Tool. medRxiv; 2021.

39. Awini E, Agyepong IA, Owiredu D, Gyimah L, Ashinyo ME, Yevoo LL, et al. Burden of mental health problems among pregnant and postpartum women in sub-Saharan Africa: systematic review and meta-analysis protocol. BMJ Open. 2023;13(6):e069545.

40. Danso-Appiah A, Owiredu D, Akuffo KO. Praziquantel-related visual disorders among recipients in mass drug administration campaigns in schistosomiasis endemic settings: Systematic review and meta-analysis protocol. PLoS One. 2024;19(5):e0300384.

41. Gyimah L, Agyepong IA, Owiredu D, Awini E, Yevoo LL, Ashinyo ME, et al. Tools for screening maternal mental health conditions in primary care settings in sub-Saharan Africa: systematic review. Frontiers in Public Health. 2024;12.

42. Danso-Appiah A, Olliaro PL, Donegan S, Sinclair D, Utzinger J. Drugs for treating Schistosoma mansoni infection. Cochrane Database Syst Rev. 2013;2013(2):Cd000528.

43. Mirzoev T, Koduah A, Cronin de Chavez A, Baatiema L, Danso-Appiah A, Ensor T, et al. Implementation of medicines pricing policies in sub-Saharan Africa: protocol for a systematic review. BMJ Open. 2021;11(2):e044293.

44. Moher D, Liberati A, Tetzlaff J, Altman DG. Preferred reporting items for systematic reviews and meta-analyses: the PRISMA statement. PLoS medicine. 2009;6(7):e1000097.

45. Page MJ, McKenzie JE, Bossuyt PM, Boutron I, Hoffmann TC, Mulrow CD, et al. The PRISMA 2020 statement: an updated guideline for reporting systematic reviews. BMJ (Clinical research ed). 2021;372:n71.

46. Ouzzani M, Hammady H, Fedorowicz Z, Elmagarmid A. Rayyan-a web and mobile app for systematic reviews. Systematic reviews. 2016;5(1):210.

47. Abdulai M, Owiredu D, Boadu I, Tabong PT, Sarfo B, Bonful HA, et al. Psychosocial interventions and their effectiveness on quality of life among elderly persons living with HIV in Africa South of the Sahara: Systematic review and meta -analysis protocol. PLoS One. 2023;18(9):e0291781.

48. Hoy D, Brooks P, Woolf A, Blyth F, March L, Bain C, et al. Assessing risk of bias in prevalence studies: modification of an existing tool and evidence of interrater agreement. Journal of clinical epidemiology. 2012;65(9):934–9.

49. United Nations. Standard country or area codes for statistical use (M49): Geographic Regions: United Nations Statistical Division; 2024 [Available from: https://unstats.un.org/unsd/methodology/m49/.

50. Higgins JP, Thompson SG, Deeks JJ, Altman DG. Measuring inconsistency in meta-analyses. BMJ (Clinical research ed). 2003;327(7414):557–60.

51. Huedo-Medina TB, Sánchez-Meca J, Marín-Martínez F, Botella J. Assessing heterogeneity in meta-analysis: Q statistic or I2 index? Psychological methods. 2006;11(2):193–206.

52. Cochrane UK. How to read a forest plot? : Cochrane UK; 2016 [updated 11th July 2022 Available from: https://uk.cochrane.org/news/how-read-forest-plot.

53. Sedgwick P. Meta-analyses: what is heterogeneity? BMJ (Clinical research ed). 2015;350:h1435.

54. Ngongo CJ, Raassen T, Lombard L, Roosmalen J, Weyers S, Temmerman M, et al. Delivery mode for prolonged, obstructed labour resulting in obstetric fistula: a retrospective review of 4396 women in East and Central Africa. BJOG: An International Journal of Obstetrics & Gynaecology. 2020;127(6):702–7.

55. Ngongo CJ, Raassen T, Mahendeka M, Lombard L, van Roosmalen J. Iatrogenic genito-urinary fistula following cesarean birth in nine sub-Saharan African countries: a retrospective review. BMC pregnancy and childbirth. 2022;22(1):541.

56. Browning A. The circumferential obstetric fistula: characteristics, management and outcomes. J BJOG: An International Journal of Obstetrics Gynaecology. 2007;114(9):1172–6.

57. Browning A, Whiteside S. Characteristics, management, and outcomes of repair of rectovaginal fistula among 1100 consecutive cases of female genital tract fistula in Ethiopia. International journal of gynaecology and obstetrics: the official organ of the International Federation of Gynaecology and Obstetrics. 2015;131(1):70–3.

58. Ezegwui H, Nwogu-Ikojo E. Vesico-vaginal fistula in Eastern Nigeria. 2005;25(6):589–91.

59. Obi SN, Ozumba BC, Onyebuchi AK. Decreasing incidence and changing aetiological factors of vesico-vaginal fistula in south-east Nigeria. Journal of obstetrics and gynaecology : the journal of the Institute of Obstetrics and Gynaecology. 2008;28(6):629–31.

60. Ijaiya MA, Aboyeji PA. Obstetric urogenital fistula: The Ilorin experience, Nigeria. West African Journal of Medicine. 2004;23(1):7–9.

61. Ijaiya M-d, A, Aboyeji AP, Ijaiya ZB. Epidemiology of vesico-vaginal fistula at the University of Ilorin Teaching Hospital, Ilorin, Nigeria. 2002;19(2):101–3.

62. Kazaura MR, Kamazima RS, Mangi EJ. Perceived causes of obstetric fistulae from rural southern Tanzania. African Health Sciences. 2011;11(3):377–82.

63. Mteta KA, Mbwambo J, Mvungi M. Iatrogenic ureteric and bladder injuries in obstetric and gynaecologic surgeries. East African medical journal. 2006;83(2):79–85.

64. Gumodoka B, Mach E, Majinge CR. Genito-urinary fistula patients at Bugando Medical Centre. East African medical journal. 2010;87(7):294–8.

65. Browning A, Mbise F, Foden P. The effect of early pregnancy on the formation of obstetric fistula. International Journal of Gynecology and Obstetrics. 2017;138(3):288–92.

66. Raassen T, Ngongo CJ, Mahendeka MM. Diagnosis and management of 365 ureteric injuries following obstetric and gynecologic surgery in resource-limited settings. International urogynecology journal. 2018;29(9):1303–9.

67. Siddle K, Vieren L, Fiander A. Characterising women with obstetric fistula and urogenital tract injuries in Tanzania. International Urogynecology Journal and Pelvic Floor Dysfunction. 2014;25(2):249–55.

68. Emmanuel O, Ifeanyichukwu D, Chinwendu A, Chijioke O, Uzoma O, Uzoma M, Okechukwu AJIJGO. Preliminary outcome of the management of vesicovaginal fistulae at a teaching hospital In Southeastern Nigeria. 2012;16(1):22–5.

69. Obarisiagbon EO, Olagbuji BN, Onuora VC, Oguike TC, Ande AB. Iatrogenic urological injuries complicating obstetric and gynaecological procedures. Singapore medical journal. 2011;52(10):738–41.

70. Egziabher TG, Eugene N, Ben K, Fredrick K. Obstetric fistula management and predictors of successful closure among women attending a public tertiary hospital in Rwanda: A retrospective review of records. BMC Research Notes. 2015;8(1).

71. Sanda G, Chipkao R, Harissou A, Soumana A, Tassiou EM. Iatrogenic genito-urinary fistulae: A report of 62 cases and literature review. African Journal of Urology. 2016;22(2):55–60.

72. Gele AA, Salad AM, Jimale LH, Kour P, Austveg B, Kumar B. Relying on Visiting Foreign Doctors for Fistula Repair: The Profile of Women Attending Fistula Repair Surgery in Somalia. Obstetrics and gynecology international. 2017;2017:6069124.

73. Meurice M, Genadry R, Heimer C, Ruffer G, Kafunjo BJ. Social Experiences of Women with Obstetric Fistula Seeking Treatment in Kampala, Uganda. Annals of Global Health. 2017;83(3-4):541–9.

74. Lewis Wall L, Belay S, Haregot T, Dukes J, Berhan E, Abreha M. A case–control study of the risk factors for obstetric fistula in Tigray, Ethiopia. International urogynecology journal. 2017;28(12):1817–24.

75. Paluku J, Bruce P, Kamabu E, Kataliko B, Kasereka J, Dube A. Childbirth-associated fistula and perineal tears repaired on outreach campaigns in remote democratic republic of congo. Int J Womens Health. 2021;13:1025–31.

76. Bernard L, Giles A, Fabiano S, Giles S, Hudgins S, Olson A, et al. Predictors of obstetric fistula repair outcomes in Lubango, Angola. 2019;41(12):1726–33.

77. Chang OH, Stokes MJ, Chalamanda C, Wilkinson J, Pope RJ. Baseline renal function and renal ultrasound findings in patients with obstetric fistulas (RENFRU): a prospective cohort study. BJOG: An International Journal of Obstetrics and Gynaecology. 2020;127(7):897–904.

78. Lawal OO, Abdus-salam RA, Bello OO, Morhason-Bello IO, Ojengbede OA. Outcome of urethral reconstruction among vesico-vaginal fistula patients: a cross-sectional study. 2021;27:1–7.

79. Sarr A, Ze Ondo C, Thiam A, Badji CA, Sine B, Ndiaye M, et al. Epidemiological, etiological and evolutionary profile of vesico-vaginal fistulas in Senegal. Progres en Urologie. 2023;33(7):401–6.

80. Vodouhe MV, Foma JdDY, Gandaho KI, Kakpo MZ, Atade SR, Sambo BT, et al. Epidemiological and Therapeutic Aspects of Obstetric Fistula in 2021: A Review of 97 Cases at the Departmental University Hospital Centre of Borgou and Alibori in Benin. 2023;13(6):1007–19.

81. Kpatcha TM, Tengue K, Adodo A, Wangala P, Botcho G, Leloua E, et al. [Ureterovaginal fistula after caesarean: Diagnosis and management in a resource-constrained hospital in Togo]. Bulletin de la Societe de pathologie exotique (1990). 2016;109(5):329–33.

82. Maroyi R, Moureau MK, Brown HW, Ajay R, Byabene G, Mukwege DM. Etiology and factors associated with urogenital fistula among women who have undergone cesarean section: a cross-sectional study. 2023;23(1).

83. Ijaiya MA, Rahman AG, Aboyeji AP, Olatinwo AWO, Esuga SA, Ogah OK, et al. Vesicovaginal fistula: A review of nigerian experience. West African Journal of Medicine. 2010;29(5):293–8.

84. El-Tabey NA, Ali-El-Dein B, Shaaban AA, El-Kappany HA, Mokhtar AA, El-Azab M, Kheir AA. Urological trauma after gynecological and obstetric surgeries: An 18-year single-center experience. Scandinavian Journal of Urology and Nephrology. 2006;40(3):225–31.

85. Capes T, Ascher-Walsh C, Rogo-Gupta L, Lo Y, Idrissa A, editors. OBSTETRIC FISTULA IN NIAMEY, NIGER: A RETROSPECTIVE REVIEW OF 700 PATIENTS. Neurourology and urodynamics; 2011: WILEY-LISS DIV JOHN WILEY & SONS INC, 111 RIVER ST, HOBOKEN, NJ 07030 USA.

86. Williams G, Broughton S, Worku H, Tekle H. Five years experience of ureterovaginal fistulae following obstetric or gynecological intervention in Ethiopia. African Journal of Urology. 2010;16(1):17–9.

87. Anemu Sori D, Workineh Azale A, Hiko Gemeda D, Sori DA, Azale AW, Gemeda DH. Characteristics and repair outcome of patients with Vesicovaginal fistula managed in Jimma University teaching Hospital, Ethiopia. BMC Urology. 2016;16:1–6.

88. Belayihun B, Mavhandu-mudzuis, Helen A. Psychological distress in women with obstetric fistula in Ethiopia: a multi-center, facility-based, cross-sectional study. Ethiopian Journal of Health Development. 2018;32(4).

89. Areba AS, Akessa GM, Tadesse M, Haile A, Abire BG, Eritero AC, et al. Recovery Time and Its Predictors among Women Admitted with Obstetric Fistula in Jimma University Medical Center Southwest, Ethiopia: A Retrospective Cohort Study. medRxiv. 2022:2022.02.02.22270299.

90. Ala SH, Husain S. Outcome of Vesicovaginal Fistula Repair: An Experience at Hamlin Fistula Hospital, Ethiopia. 2022;14(1):11–3.

91. Meikena KH, Bihon AM, Serka S. Predictors and outcomes of surgical repair of obstetric fistula at Mekelle Hamlin Fistula Center, Northern Ethiopia. 2023;34(8):1891–8.

92. Tadesse S, Ejigu N, Edosa D, Ashegu T, Dulla D. Obstetric fistula repair failure and its associated factors among women underwent repair in Yirgalem Hamlin fistula center, Sidama Regional State, Southern Ethiopia, 2021: a retrospective cross sectional study. 2022;22(1):288.

93. Hawkins L, Spitzer RF, Christoffersen-Deb A, Leah J, Mabeya H. Characteristics and surgical success of patients presenting for repair of obstetric fistula in western Kenya. International journal of gynaecology and obstetrics: the official organ of the International Federation of Gynaecology and Obstetrics. 2013;120(2):178–82.

94. Mwangi HR. Factors associated with obstetric fistula repair failure among women admitted at Gynocare Women’s and fistula Hospital in Kenya, 2012-2016: a case control study: University of Nairobi; 2017.

95. Obi S, Ozumba B, Onyebuchi A. Decreasing incidence and changing aetiological factors of vesico-vaginal fistula in south-east Nigeria. 2008;28(6):629–31.

96. Emasu A, Ruder B, Wall LL, Matovu A, Alia G, Barageine JK. Reintegration needs of young women following genitourinary fistula surgery in Uganda. International urogynecology journal. 2019;30(7):1101–10.

97. Kayondo M, Wasswa S, Kabakyenga J, Mukiibi N, Senkungu J, Stenson A, Mukasa P. Predictors and outcome of surgical repair of obstetric fistula at a regional referral hospital, Mbarara, western Uganda. 2011;11:1–9.

98. Sebukoto HR, Semwaga E, Rugakingila RA. Urological injuries following obstetrical and gynecological surgeries. East and Central African Journal of Surgery. 2016;21:148.

99. Cichowitz C, Watt MH, Mchome B, Masenga GG. Delays contributing to the development and repair of obstetric fistula in northern Tanzania. 2018;29(3):397–405.

100. Lilungulu A, Gumodoka B, Nassoro M, Soka P, Stephen K. Obstetric fistulae, birth out comes, and sur gical repair outcomes: a retrospective analysis of hospital-based data in Dodoma, Tanzania. 2018.

101. Richter LA, Lee H, Nishimwe A, Niteka LC, Kielb SJ. Characteristics of Genitourinary Fistula in Kigali, Rwanda; 5-Year Trends. Urology. 2021;150:165–9.

102. Washington BB, Raker CA, Kabeja GA, Kay A, Hampton BS. Demographic and delivery characteristics associated with obstetric fistula in Kigali, Rwanda. International Journal of Gynecology and Obstetrics. 2015;129(1):34–7.

103. Hategekimana T, Rwamasirabo E, Banamwana R, Van den Ende J. Results and predictions of success of vesico-vaginal fistula repair at a national reference level in RwandaRésultats et facteurs prédicateurs du résultat chirurgical des fistules vésico-vaginales à l’échelle d’un hôpital de reference national. African journal of urology. 2005;11(4):261–7.

104. Holme A, Breen M, MacArthur C. Obstetric fistulae: a study of women managed at the Monze Mission Hospital, Zambia. Bjog. 2007;114(8):1010–7.

105. Nambala N. A study to determine women’s intention to prevent Vesico-Vaginal Fistula recurrence in two repair centres in Zambia 2012.

106. Imakando MM, Michelo C, Mkandawire T, Kasonka L. Characteristics and Surgical Repair Outcomes of Obstetric Fistula Patients Managed at a Teaching Hospital in Zambia: A Retrospective Cross Sectional Study. J Medical Journal of Zambia. 2022;49(2):146–56.

107. Yaay JKA, Athian AA, Akoon DDT, Fabiano AA, Ariath TB, Mona AA. Risk factors for vesicovaginal and rectovaginal fistulae in women treated at Juba Teaching Hospital in 2020-2021: A retrospective study. 2022;15(2):54–7.

108. Sereke D, Hailemelecot H, Issak Y, Estifanose DJS. Obstetric Vesico-vaginal Fistulae: a documentary review of women managed in Mendefera Zonal Referral and National Fistula Hospital, Eritrea. 2020;8(5):149–54.

109. Mocumbi S, Hanson C, Högberg U, Boene H, von Dadelszen P, Bergström A, et al. Obstetric fistulae in southern Mozambique: incidence, obstetric characteristics and treatment. Reproductive health. 2017;14(1):147.

110. Raassen TJ, Verdaasdonk EG, Vierhout ME. Prospective results after first-time surgery for obstetric fistulas in East African women. Int Urogynecol J Pelvic Floor Dysfunct. 2008;19(1):73–9.

111. Wall LL, Karshima JA, Kirschner C, Arrowsmith SD. The obstetric vesicovaginal fistula: characteristics of 899 patients from Jos, Nigeria. 2004;190(4):1011–6.

112. Kirschner CV, Yost KJ, Du H, Karshima JA, Arrowsmith SD, Wall LL. Obstetric fistula: the ECWA Evangel VVF Center surgical experience from Jos, Nigeria. 2010;21:1525–33.

113. Olusegun AK, Akinfolarin AC, Olabisi LM. A review of clinical pattern and outcome of vesicovaginal fistula. Journal of the National Medical Association. 2009;101(6):593–5.

114. Daru P, Karshima J, Mikah S, Nyango D. The burden of vesico-vaginal fistula in North Central Nigeria. 2011;1(2):50.

115. Ojewola RW, Tijani KH, Jeje EA, Ogunjimi MA, Animashaun EA, Akanmu ON. Transabdominal repair of vesicovaginal fistulae: A 10-year tertiary care hospital experience in Nigeria. 2018;25(4):213–9.

116. Sunday-Adeoye I, Okonta P, Ulu OL. Prevalence, profile and obstetric experience of fistula patients in Abakaliki, Southeast Nigeria. 2011;25(1):e6.

117. Ngwan SD, Edem BE, Anzaku AS, Eke BA, Shittu MA, Obekpa SA. Vesico-vaginal fistula repair: experience with first twenty-three patients seen at a tertiary hospital in north-central Nigeria. 2015;15(2):66–8.

118. Nasir S, Elladan A, Hassan M, Panti A. Pattern and Outcome of Iatrogenic Genitourinary Fistula from Obstetric and Gynaecological Surgeries in a Tertiary Institution, North-Western Nigeria. Asian Journal of Medicine and Health. 2018;10(3):1–7.

119. Abah MG, Lengmang SJ, Inyang-Etoh EC, Abah I. Socio-Demographic and Obstetric Profile of Women Who Presented for Obstetric Fistula Repair in the Middle Belt Region of Nigeria. Journal of Womens Health Development. 2022;5(1):15–23.

120. Lawal O, Bello O, Morhason-Bello I, Abdus-Salam R, Ojengbede O. Our Experience with Iatrogenic Ureteric Injuries among Women Presenting to University College Hospital, Ibadan: A Call to Action on Trigger Factors. 2019;2019:6456141.

121. Ekanem EI, Ekott MI, Ekabua JE, Agan TU, Inyang-Otu A. Outcome of management of obstetric genito-urinary fistulae in the General Hospital, Ikot Ekpene, Akwa Ibom state, Nigeria. 2010;24(2):e1.

122. Kabore FA, Nama SDA, Ouedraogo B, Kabore M, Ouattara A, Kirakoya B, Karsenty G. Characteristics of Obstetric and Iatrogenic Urogenital Fistulas in Burkina Faso: A Cross-Sectional Study. Advances in Urology. 2021:1–7.

123. Traore TM, Ouedraogo S, Kabore M, Ouedraogo S, Traore JJ. Characteristics of obstetric urogenital fistulas in a regional teaching hospital in Burkina Faso: a retrospective cross-sectional study. Pan African Medical Journal. 2023;44.

124. Ouedraogo I, Payne C, Nardos R, Adelman AJ, Wall LL. Obstetric fistula in Niger: 6-month postoperative follow-up of 384 patients from the Danja Fistula Center. International urogynecology journal. 2018;29(3):345–51.

125. Munoz O, Bowling CB, Gerten KA, Taryor R, Norman AM, Szychowski JM, et al. Factors influencing post-operative short-term outcomes of vesicovaginal fistula repairs in a community hospital in Liberia. 2011;4(6):259–65.

126. Sangaré D, Berthé HJG, Diakité ML, Samassekou A, Diakité AS, Diallo MS, et al. [Urological Complications of Pelvic Surgery at Point-G Hospital, About 23 Cases]. Le Mali medical. 2018;33(2):9–12.

127. Sheku MG, Kabia AB, Gegbe F, Bangura EA, Fornah F, Kargbo AB, et al. A Situational Analysis of Vesico-Vaginal Fistula at the Aberdeen Women’s Centre-Freetown. 2021;5(7):12.

128. Kpatcha TM, Wangala P, Botcho G, Tchandana M, Nembuzu D, Aboubakari AS. Epidemiologic, anatomoclinic and therapeutic profil of urogenital and rectovaginal fistula in TOGO. Progres en Urologie. 2020;30(11):597–603.

129. Mafu MM, Banze DFK, Aussak BTT, Kolié D, Camara BS, Nembunzu D, et al. Factors associated with surgical repair success of female genital fistula in the Democratic Republic of Congo: Experiences of the Fistula Care Plus Project, 2017-2019. Tropical Medicine & International Health. 2022;27(9):831–9.

130. Loposso MN, Ndundu J, De Win G, Ost D, Punga AM, De Ridder D. Obstetric fistula in a district hospital in DR Congo: Fistula still occur despite access to caesarean section. Neurourology and urodynamics. 2015;34(5):434–7.

131. Sjøveian S, Vangen S, Mukwege D, Onsrud M. Surgical outcome of obstetric fistula: a retrospective analysis of 595 patients. Acta obstetricia et gynecologica Scandinavica. 2011;90(7):753–60.

132. Nsambi JB, Mukuku O, Kakudji PL, Kakoma J-BS. Socio-demographic and delivery characteristics of patients with obstetric fistula in Haut-Katanga Province, Democratic Republic of Congo. 2019;7(3):1–5.

133. Paluku JL, Sikakulya FK, Furaha CM, Kamabu EM, Aksanti BK, Tsongo ZK, et al. Epidemiological, anatomoclinical, and therapeutic profile of obstetric fistula in the Democratic Republic of the Congo: About 1267 patients. J Tropical medicine & international health 2024.

134. Tebeu PM, Maninzou SD, Kengne Fosso G, Jemea B, Fomulu JN, Rochat CH. Risk factors for obstetric vesicovaginal fistula at University Teaching Hospital, Yaoundé, Cameroon. International journal of gynaecology and obstetrics: the official organ of the International Federation of Gynaecology and Obstetrics. 2012;118(3):256–8.

135. Bouya PA, Itoua Nganongo W, Lomina D, Iloki LH. Retrospective study of 34 obstetrical uro-genital fistulas. Gynecologie Obstetrique et Fertilite. 2002;30(10):780–3.

136. Naidoo TD, Moodley J, Naidoo S. Genital tract fistula: a case series from a tertiary centre in South Africa. International urogynecology journal. 2018;29(3):383–9.

137. Ramphal S, Kalane G, Fourie T, Moodley J. Obstetric urinary fistulas in KwaZulu-Natal - what is the extent of this tragedy? South African Medical Journal. 2007;13:92–6.

138. Federspiel F, Mukhopadhyay S, Milsom PJ, Scott JW, Riesel JN, Meara JG. Global surgical, obstetric, and anesthetic task shifting: A systematic literature review. Surgery. 2018;164(3):553–8.

139. Betran AP, Ye J, Moller AB, Souza JP, Zhang J. Trends and projections of caesarean section rates: global and regional estimates. BMJ Glob Health. 2021;6(6).

140. López-Carpintero N, de la Fuente-Valero J, Salazar-Arquero FJ, Hernández-Aguado JJ. [Great vesicovaginal fistula after normal vaginal delivery in a developed country]. Ginecol Obstet Mex. 2015;83(12):798–802.

141. Reisenauer C. Presentation and management of vesicovaginal fistulae after delivery at a German women’s hospital. Arch Gynecol Obstet. 2017;296(1):1–3.

142. Bergström S. Training non-physician mid-level providers of care (associate clinicians) to perform caesarean sections in low-income countries. Best Pract Res Clin Obstet Gynaecol. 2015;29(8):1092–101.

143. van Duinen AJ, Kamara MM, Hagander L, Ashley T, Koroma AP, Leather A, et al. Caesarean section performed by medical doctors and associate clinicians in Sierra Leone. The British journal of surgery. 2019;106(2):e129–e37.

144. KMPDC. Internship Requirements Kenya: Kenya Medical Practitioners and Dentists Council; 2024 [Available from: https://kmpdc.go.ke/internship/.

145. Ramoolla B, Van der Haar G, Luke A, King R, Jacob N, Luke B. Medical internship training in South Africa: Reflections on the new training model 2020-2021. South African Medical Journal. 2023;113(5):1195–8.

146. Miller AC, Mayanja F, Porter JD. Assessing South African medical interns’ experience and confidence in managing obstetric emergencies. S Afr Med J. 2021;111(11):1098–103.

147. Wehrmeister FC, Fayé CM, da Silva ICM, Amouzou A, Ferreira LZ, Jiwani SS, et al. Wealth-related inequalities in the coverage of reproductive, maternal, newborn and child health interventions in 36 countries in the African Region. Bull World Health Organ. 2020;98(6):394–405.

148. Sidze EM, Wekesah FM, Kisia L, Abajobir A. Inequalities in Access and Utilization of Maternal, Newborn and Child Health Services in sub-Saharan Africa: A Special Focus on Urban Settings. Matern Child Health J. 2022;26(2):250–79.

149. Garces A, McClure EM, Espinoza L, Saleem S, Figueroa L, Bucher S, Goldenberg RL. Traditional birth attendants and birth outcomes in low-middle income countries: A review. Semin Perinatol. 2019;43(5):247–51.

150. Souza JP, Day LT, Rezende-Gomes AC, Zhang J, Mori R, Baguiya A, et al. A global analysis of the determinants of maternal health and transitions in maternal mortality. Lancet Glob Health. 2024;12(2):e306–e16.

151. Ameh CA, Kerr R, Madaj B, Mdegela M, Kana T, Jones S, et al. Knowledge and Skills of Healthcare Providers in Sub-Saharan Africa and Asia before and after Competency-Based Training in Emergency Obstetric and Early Newborn Care. PloS one. 2016;11(12):e0167270.

152. Sehar N, Azam A, Ara S, Zaman F, Sarkar N, Hyder F. Iatrogenic Fistula after Gynecological Operation in a Tertiary level Hospital. Central Medical College Journal. 2023;6:28–33.

153. Wilson A, Lissauer D, Thangaratinam S, Khan KS, MacArthur C, Coomarasamy A. A comparison of clinical officers with medical doctors on outcomes of caesarean section in the developing world: meta-analysis of controlled studies. BMJ (Clinical research ed). 2011;342:d2600.

154. Ngongo CJ, Raassen T, van Roosmalen J, Mahendeka M, Lombard L, Bukusi E. Equivalence between physicians and associate clinicians in the frequency of iatrogenic urogenital fistula following cesarean section in Tanzania and Malawi. Hum Resour Health. 2024;22(1):43.

